# T cell plasticity in systemic lupus erythematosus revealed by large-scale T cell receptor repertoire and transcriptome studies

**DOI:** 10.1101/2025.01.06.24319648

**Authors:** Yasuo Nagafuchi, Masahiro Nakano, Kaitlyn A. Lagattuta, Mineto Ota, Hiroaki Hatano, Haruka Takahashi, Takahiro Itamiya, Hajime Inokuchi, Soumya Raychaudhuri, Tomohisa Okamura, Keishi Fujio, Kazuyoshi Ishigaki

## Abstract

CD4+ T cell plasticity plays a pivotal role in immune homeostasis. However, evidence of T cell plasticity and its pathological role in human systemic lupus erythematosus (SLE) is missing due to the lack of a reporter system. Here we utilized T cell receptor (TCR) repertoire data as a molecular signatures alongside transcriptomic dataset. Using a large-scale ImmuNexUT database of autoimmune disease patients including 117 SLE cases, we quantified T cell plasticity across 13 fine-grained T cell-types. We analyzed 6,392 samples in total and identified two orthogonal signatures of repertoire and transcriptome, the cell-type and disease signatures, allowing us to investigate CD4+ T cell plasticity comprehensively. Among all possible patterns, the plasticity level was the highest in effector regulatory T cells (eTreg) to Th1 plasticity, which was replicated in an independent cohort. Moreover, eTreg-to-Th1 plasticity positively correlated with SLE disease activity. Our study provides novel evidence that Treg plasticity is involved in SLE pathology.

## Introduction

Systemic lupus erythematosus (SLE) is a chronic autoimmune disease marked by a myriad of clinical symptoms and autoantibodies^1^. Many lines of evidence support the important role of CD4+ T cell in SLE pathogenesis. CD4+ T cells guide B cells to produce autoantibodies ^2, 3^, infiltrate into multiple tissues^3, 4, 5^, show disrupted abundance in peripheral blood^2, 3, 6, 7, 8^, and produce dysregulated inflammatory cytokines^7, 9^. Human CD4+ T cells include transcriptionally and functionally distinct cell types, such as Th1, Th2, Th17, follicular helper cells, and regulatory T cells (Treg cells)^10, 11^. In addition to resting Treg cells (Fr. I) and activated Treg cells (Fr. II), a subset of activated Treg-like cells with low *FOXP3* expression levels (named Fr. III) has been described^6^. Fr. III T cells are not immunosuppressive and produce more IL-17 than other CD4+ T cell-types^6^.

Intriguingly, the transcriptional state of each T cell is not fixed but rather plastic. For example, Treg cells transition into effector Th17 or Th1 cells in mice under experimental inflammatory conditions^12, 13^. This shift involves the loss of the characteristic Treg *FOXP3* expression and immunosuppressive function. In untreated multiple sclerosis patients, an increase in Th1-like, interferon-γ (IFN-γ)-secreting FOXP3(+) T cells with reduced in vitro suppressive activity has been reported^14^. Nonetheless, there is scarce data about Treg plasticity in SLE patients.

The largest obstacle to investigating human T cell plasticity is the lack of a reporter system that can track previous cellular identities, such as FOXP3-GFP marker in mice^12, 13^. However, when a T cell proliferates, its unique T cell antigen receptor (TCR) nucleotide sequence is replicated in daughter cells. Thus, the TCR sequence, a product of stochastic gene recombination, is a unique molecular “fingerprint” that can track T cells and their progeny through clonal proliferation. We and others have observed that T cell-types have characteristic TCR sequence patterns; for example, CDR3 amino acid hydrophobicity is a major distinguishing characteristic of Tregs^15^. This TCR feature is likely a causal factor towards Treg fate, given the stochastic nature of TCR gene recombination prior to Treg differentiation. Analogously, CD4+ T cells and CD8+ T cells demonstrate distinct TCR sequence characteristics^16^ ^17^. Although T cell types are typically inferred through transcriptional data, the T cell transcriptome is not inherently constant due to T cell plasticity. Therefore, TCR and transcriptome data together provide complementary evidence of previous T cell states.

In this study, we sought to obtain a global picture of the T cell plasticity in SLE patients. To do so, we revisited the ImmuNexUT (Immune Cell Gene Expression Atlas from the University of Tokyo) database: a large-scale RNA-seq database of fine-grained immune cell types, including nine cell types for CD4+ T cells and four for CD8+ T cells. These data were collected from SLE patients (n=117), nine other immune-mediated disease (IMD; n=317) patients, and healthy controls (HC; n=134)^18, 19^. With 6,392 samples, we quantified TCR sequence overlap between T cell types, identified TCR profiles for each T cell type, and examined how these results differed in SLE cases compared to healthy controls (**Figure 1**). For pairs of T cell-types with evidence of plasticity by TCR sequence overlap, we asked whether SLE patients exhibited the TCR characteristics of type A in cell type B or vice versa. These analyses revealed that SLE Th1 cells harbor TCR features that are typically characteristic of Tregs. Because stochastic TCR recombination precedes T cell transcriptional fate, the presence of Treg TCR features in SLE Th1 cells implies directional plasticity from Treg to Th1. We then tested the replicability of our findings in two independent cohorts and evaluated the clinical relevance of the plasticity. Our study provided a comprehensive picture of T cell-type plasticity from the combined viewpoint of TCR and transcriptome data at an unprecedented scale.

**Figure 1.**
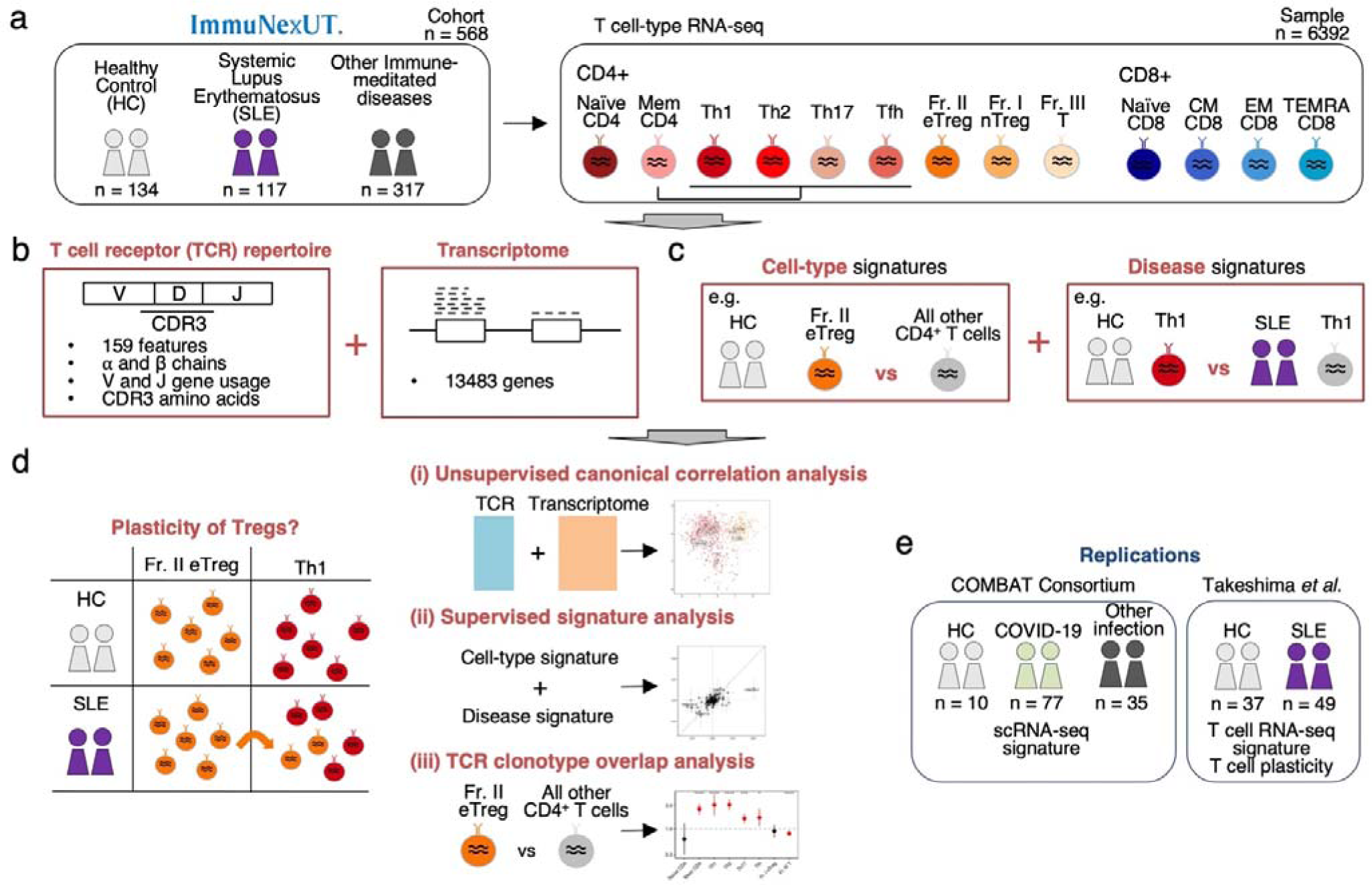
Study overview. (a) We recruited 6,392 T cell RNA-seq samples from 568 individuals, including 134 healthy controls (HC) and 117 systemic lupus erythematosus (SLE) patients and (b) performed T cell receptor repertoire (TCR) and transcriptome analysis. In the TCR analysis, we selected 159 TCR features encompassing both V and J gene usage and CDR3 region amino acid usages. (c) We characterized the cell-type and disease signatures. Cell-type signatures are specific to each T cell-type and were calculated using only HC samples. Disease signatures are case-control signatures of the SLE patients in each T cell-type. (d) We aimed to investigate the plasticity of CD4+ T cells through three approaches: (i) unsupervised canonical correlation analysis of TCR and transcriptome datasets, (ii) supervised correlation analysis of the two identified signatures, and (iii) TCR clonotype overlap analysis. (e) The signatures and T cell plasticity were externally validated using public datasets. CDR3, complementary determining region 3; scRNA-seq, single cell RNA-seq. T cell-type abbreviations are summarized in Supplementary Table 2.

## Results

### Covariations between TCR and transcriptome datasets supported cell type plasticity in IMD patients

ImmuNexUT has 117 patients with SLE, 317 patients with nine other IMDs, and 134 HC (n=568 in total; **Supplementary Table 1**). We sorted nine CD4+ T cell-types and four CD8+ T cell-types, targeting 5,000 cells for each sample, and conducted RNA-seq (**Figure 1a**, **Supplementary Table 2**). In sorting the effector CD4+ T cell-types, CD25+ regulatory T cells (Fr. I and Fr. II) and Treg-like Fr. III cells were excluded, while the remaining memory and effector CD4+ T cells were designated as “Mem CD4.” These cells were further subdivided into Th1, Th2, Th17, and Tfh cells based on canonical surface markers: CXCR3 for Th1, CCR4 for Th2, CCR6 for Th17, and CXCR5 for Tfh. Following quality controls, 6,392 samples from 568 donors remained in total, and we prepared TCR and transcriptome data for the downstream analyses (see details in the **Supplementary Note**).

We extracted RNA-seq reads covering TCR mRNA and successfully identified TCR amino acid sequences from each sample. On average, we found 876 and 1,034 distinct functional clonotypes from alpha and beta chains, respectively. Considering that we sorted at most 5,000 cells (a.k.a., the theoretical maximum number of unique clonotypes assuming no clonal expansion), our pipeline has an efficient coverage of the TCR repertoire. From alpha and beta chains, we extracted 159 TCR repertoire sequence features: 33 CDR3 middle position amino acid frequencies, and 69 V and 57 J gene usage frequencies^20^ (**Figure 1b, Methods**).

We recently reported a substantial covariation between a paired single-cell TCR and transcriptome datasets^20^, and we first sought to replicate these findings in our bulk datasets. We tested whether TCR and transcriptome data captured the shared signatures separating cell types (**Figure 2a-d**). For this aim, we conducted regularized canonical correlation analysis (CCA) using a paired dataset of TCR features and transcriptome, following the approach in our previous study^20^. CCA is a dimension-reduction method that quantifies the covariation between “two” datasets and accordingly projects each dataset into low-dimensional space: i.e., canonical variates (CVs). CCA is clearly different from PCA, another popular dimension-reduction strategy, quantifying the variation of a “single” dataset. In this “inter-cell-type” CCA, we only included HC samples to avoid any influences from IMD disease status (1,366 samples from 134 donors). We conducted CCA in three conditions, including all T cells, CD4+ T cell, or CD8+ T cell samples. Strikingly, CCA found substantial covariations between TCR and transcriptome datasets: the canonical correlations between the CV1 pair were 0.99, 0.85, and 0.94 for all T cells, CD4+ T cell, and CD8+ T cell samples respectively (**Figure 2e-f**). Permutation tests confirmed the empirical significance of each canonical correlation (*P*_perm_ < 0.01, **Supplementary** Figure 1a, **Methods**). Compared with the PC scores calculated by TCR or transcriptome data separately, the CVs offered clear advantages for separating T cell-types. In TCR data analysis, CVs yielded much finer separation of cell-types than PCs, suggesting that TCR data alone captures some cell-type information but benefits from additional transcriptome data reflecting comprehensive cellular biological processes (**Figure 2b-d** and **Supplementary** Figure 1). Similarly, transcriptome data also benefited from integrating TCR information. As a notable example, the separation of Th2 was much more distinct in CVs than in PCs, suggesting that TCR data augmented cell-type signatures that are hard to purify using transcriptome data alone. We provide further investigation on the biological processes inferred from CCA and PCA in the **Supplementary Note**. Since CCA outputs CVs for both TCR and transcriptome, we used the CCA results of TCR datasets hereafter.

**Figure 2.**
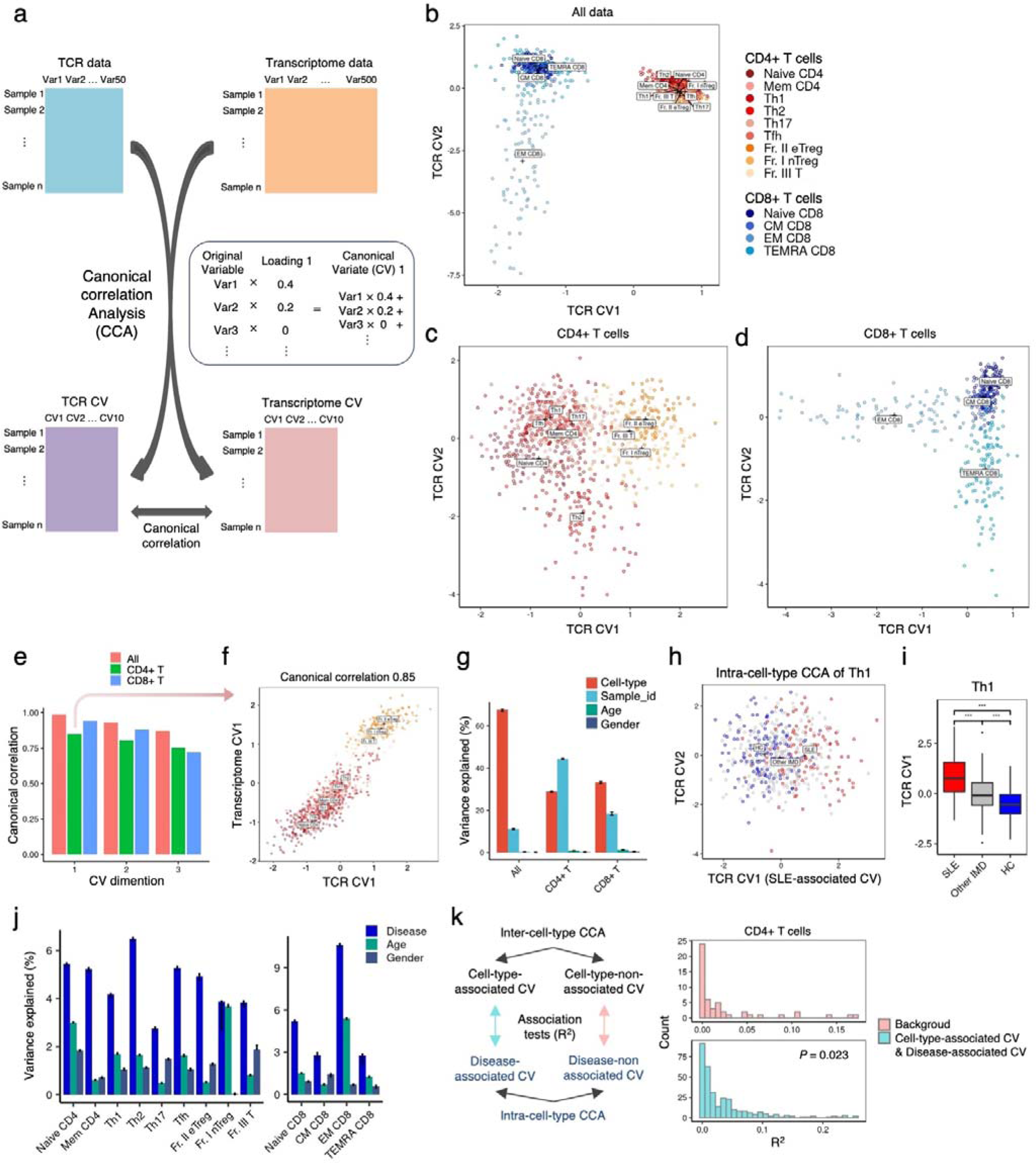
Canonical correlation analysis of the TCR and transcriptome datasets. (a) Schematic representation of canonical correlation analysis (CCA). CCA seeks to re-express the T cell receptor (TCR) and transcriptome dataset as pairs of canonical variates (CV) that are highly correlated with each other. The resulting CV is composed of the weighted sum of the original variables by the loading vectors. (b-d) Inter-cell-type CCA of TCR and transcriptome datasets was performed using all healthy control samples (b), and separately for CD4+ T cell (c) and CD8+ T cell (d) samples. The first two TCR CVs of the samples are depicted. (e) Canonical correlation of the first three TCR and transcriptome CVs. (f) Representative canonical correlation of TCR CV1 and transcriptome CV1. (g) Weighted variance partitioning analysis of the inter-cell-type CCA. The explained variance under each of the three analysis conditions is depicted. Error bars indicate the 95% confidence interval of the jackknife resampling. (h-i) Representative intra-cell-type CCA of TCR and transcriptome datasets using Th1 samples. CV1 was the SLE-associated CV. ***; *P*-value < 0.001. (j) Weighted variance partitioning analysis of the intra-cell-type CCA. The explained variance under each of the three analysis conditions is depicted. Error bars indicate the 95% confidence interval of the jackknife resampling. (k) Comparison of the association R^2^ between loading vectors of cell-type-associated inter-cell-type CVs and disease-associated intra-cell-type CVs, and cell-type-non-associated inter-cell-type CVs and disease-non-associated intra-cell-type CVs (Background). The difference was tested with Mann-Whitney U test.

To quantify the separation of the T cell-types in the CCA space, we applied weighted variance partitioning analysis (**Methods**). Cell-type labels explained substantial proportion of the overall variance in CCA: 67.5, 28.9, and 33.2 % for all T cells, CD4+ T cell, and CD8+ T cell samples (**Figure 2g**). To interpret the immunological relevance of each CC component, we tested each CC’s association with the cell-type labels.

Among CD4+ T cells or CD8+ T cells inter-cell-type CVs (n= 2 conditions x 10 CVs, 20 in total), 14 CVs were significantly associated with cell-type labels and termed them cell-type-associated CVs (n= 13 cell-types x 10 CVs, 130 tests in total, FDR < 0.05 in at least one cell-type, **Methods**). These results demonstrated that TCR and transcriptome datasets possess shared cell-type signatures, mutually complementing each other.

We then conducted “intra-cell-type” CCA to ask whether TCR and transcriptome data captured the shared signature separating IMD cases and HCs within each of the 13 cell-types, such as Th1 (**Figure 2h-i**). This is a challenging question since the variation size of intra-cell-type heterogeneity is around 78 times smaller than that of inter-cell-type heterogeneity (**Methods**). However, we successfully detected substantial correlations between TCR and transcriptome datasets in this analysis with median CV1 canonical correlation of 0.74 (ranging from 0.65 to 0.92). Disease status explained a substantial proportion of the overall variance in CCA: 4.9% in the average of T cell-types (**Figure 2j**). We then tested each CV’s association with the disease status. Among all intra-cell-type CVs (n= 13 cell-types x 10 CVs, 130 in total), 55 CVs were significantly associated with IMD status and termed them disease-associated CVs (n= 6 IMDs x 13 cell-types x 10 CVs + 4 IMDs x 8 cell-types x 10 CVs, 1,100 tests in total, FDR < 0.05 in at least one IMD, **Methods**).

The above CCA identified the shared TCR and transcriptome patterns reflecting cell-type and disease heterogeneities. We hypothesized that if T cell plasticity happens in IMD, we should observe non-zero correlations between the cell-type and disease signatures inferred by CCA. This is because, if plasticity from cell-type A to cell-type B is particular to the IMD, the disease signature of cell-type B will be contaminated by the cell-type signature of cell-type A (**Figure 1d**). To test this hypothesis, we evaluated correlations between all cell-type-associated CVs and disease-associated CVs within CD4+ T and CD8+ T cells. Compared with the background estimated non-significant CVs (**Methods**), we observed significantly larger correlation only in CD4+ T cells (**Figure 2k** and **Supplementary** Figure 1i). Therefore, we focused on the CD4+ T cell signatures hereafter.

### Cell-type signatures of TCR features and transcriptome separating CD4+ T cell-type

We next took a supervised approach to infer cell-type signatures of TCR features and transcriptome in CD4+ T cells. As in the CCA, we only included HC samples to avoid any influences from IMD disease status. In both TCR and transcriptome analyses, we applied a linear mixed model analysis of specific T cell-types, such as Fr. II eTreg, compared to all other CD4+ T cell-types, while conditioning for the effects of age, sex, and donor batch effects. This model showed improved power compared with the conventional linear models, and the permutation test confirmed well-calibrated *P*-values (**Supplementary** Figure 2a-f).

In the TCR feature analysis, we found all TCR features associated with at least one cell-type: 81 features for each cell-type on average and all 159 features for at least one cell-type (n= 159 feature x 13 cell- type, 2,067 tests in total, FDR < 0.05; **Figure 3a**; **Supplementary Table 3**). The results were well replicated in an independent dataset we reported recently^21^ (**Supplementary** Figure 3a-e). In alignment with inter-cell-type CCA that captured the Treg signatures on the CV1 axis and the Th2 signatures on the CV2 axis, we observed many TCR features associated with Tregs and Th2. For example, CDR3 β chains of the Fr. II eTreg showed higher frequency of hydrophobic amino acids than other CD4+ T cell-types, consistent with our previous report^15^ (**Figure 3b and 3e-f**). We further confirmed the increased hydrophobicity also in α chains. In Th2 analysis, basic amino acids were more preferentially used than other effector cell-types, whereas vice versa for acidic amino acids (**Figure 3c-3e, 3g**; **Supplementary** Figure 4). Interestingly, Th2 also showed the increased CDR3 hydrophobicity of CDR3 (**Figure 3c and 3e-f**). These results suggest that the physicochemical amino acid features of TCR CDR3, shaped through random recombination during thymic development, likely affects a T cell’s propensity for Treg and Th2 differentiation.

**Figure 3.**
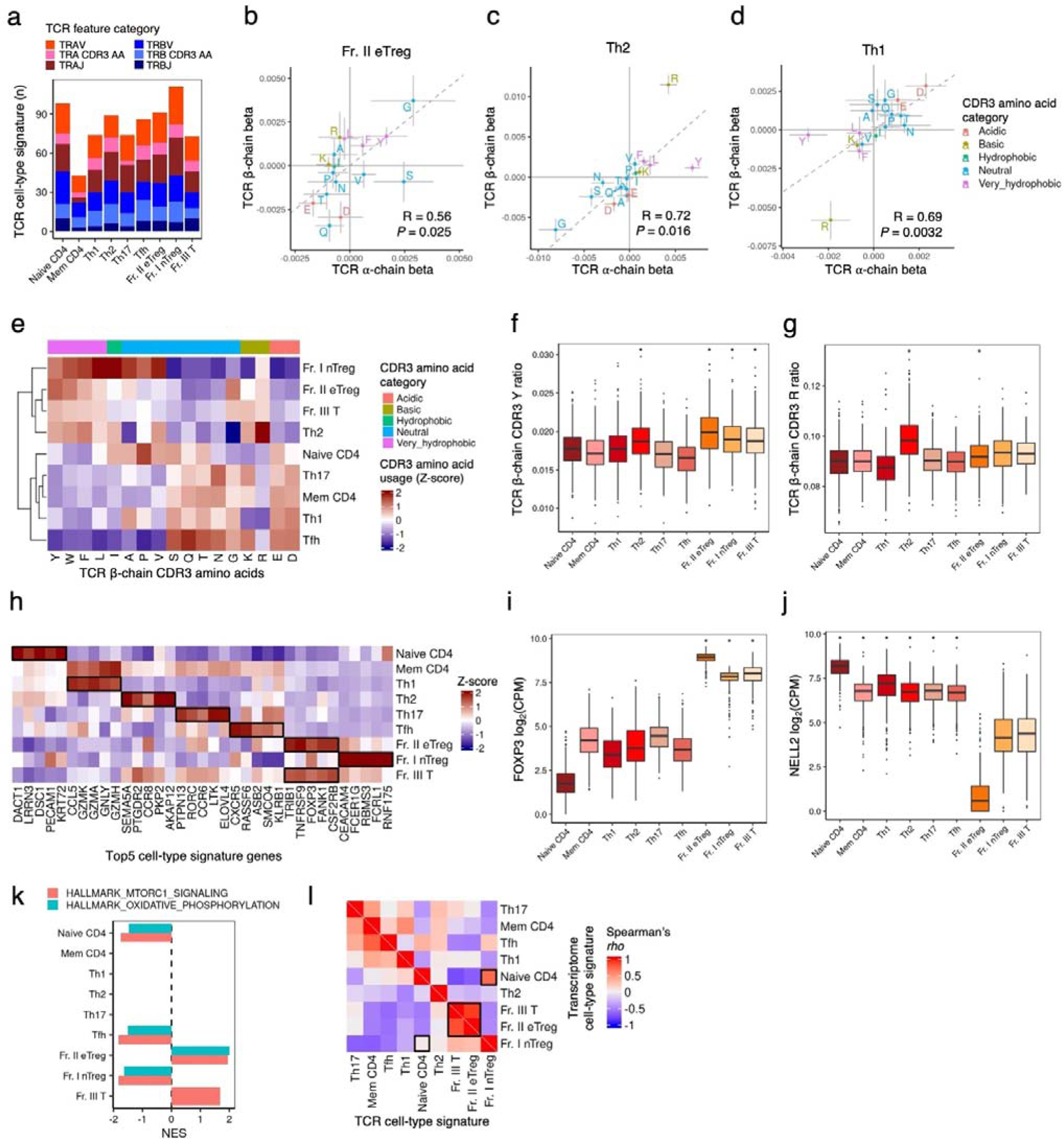
Cell-type signatures of T cell-types. (a) The number of significant cell-type CD4+ TCR signatures in T cell-types (at FDR < 0.05). TCR features were classified based on distinctions between TCR α and β chains and variations in the category (V and J gene usages and CDR3 amino acid usages). (b-d) TCR CDR3 amino acid usage signatures of T cell-types. Amino acids were categorized based on their physicochemical features, and the signature betas were compared between TCR α and β chains with Pearson Correlation Coefficient. (e) Heamap representation of TCR β-chain CDR3 amino acids usages in different CD4+ T cell-types. Each amino acid usage was normalized, and Z scores were hierarchically clustered by T cell-types. (f-g) Representative boxplots of TCR CDR3 amino acid usage in β chains. * indicates FDR < 0.05 in cell-type signature analysis. (h) Heatmap representation of the strongest cell-type signature genes in different CD4+ T cell-types. Gene expression of the top 5 genes was normalized, and Z scores were shown. The top genes of Memory CD4 T cells, which are a mixture of effector CD4+ T cell-types, were excluded. (i-k) Representative boxplots of *FOXP3* and *NELL2* genes. P-values were derived from cell-type signature analysis. (k) GSEA of cell-type signatures. Normalized enrichment score (NES) of selected Fr. II eTreg related pathways is shown. (l) A heatmap showing the Spearman correlations across CD4+ T cell-types in TCR and transcriptomic cell-type signatures. The order of T cell-type in row and column is the same and based on the hierarchical clustering using the Spearman correlation coefficients of TCR signatures. TCR, T cell receptor; FDR, False Discovery Rate; CPM, counts per million; SE, Standard error.

In transcriptome analysis, we identified 9,753 transcriptome signature genes for each T cell-type on average (n= 13,483 genes x 13 cell-type, 175,279 tests in total, 72% of the tested genes at FDR < 0.05; **Supplementary** Figure 2g; **Supplementary Table 4**). Again, we confirmed an excellent replicability in an independent cohort (**Supplementary** Figure 3a-d). This analysis validated our sorting strategy: the expression of well-established marker genes was mostly restricted to the corresponding cell-types: e.g., *CCL5* and *GZMK* for Th1, *SEMA5A* and *PTGDR2* for Th2, *RORC* and *CCR6* for Th17, and *CXCR5* and *RASSF6* for Tfh, and *FOXP3*, *TRIB1*, and *TNFRSF9* for Treg cell-types, aligning with prior literature findings^22, 23^ (**Figure 3h-i**). This analysis also identified differentially expressed genes separating closely related cell-types: e.g., down-regulated *NELL2* differentiated Fr. II eTreg from the other two Treg or Treg-like cell-types (**Figure 3j**). To explore the dysregulated biological processes in SLE T cells, we conducted Gene Set Enrichment Analysis (GSEA). This analysis shows that transcriptional pathways, such as oxidatative phosphorylation and MTORC1 signaling are upregulated in Fr. Il eTreg, in alignment with literature findings^24, 25^ (**Figure 3k** and **Supplementary** Figure 5).

We finally sought to confirm that TCR and transcriptome cell-type signatures reflected common inter-cell-type similarities, as we demonstrated in CCA. For this aim, we assessed the inter-cell-type correlation of both signatures (**Figure 3l**). Globally, both signatures showed similar correlation patterns. For example, the signatures of Fr. III T showed a tight correlation with that of Fr. II eTreg: *Rho* = 0.83, *P* < 2.2 × 10^-16^ in TCR analysis and *Rho* = 0.90, *P* < 2.2 × 10^-16^ in transcriptome. However, we also noted several distinct correlation patterns. A notable example is that Fr. I nTreg, a subpopulation of Tregs possessing a naïve phenotype. Fr. I nTreg shared transcriptome signatures with naïve CD4+ and more than other Treg or Treg-like cell-types. In contrast, Fr. I nTreg clustered with other Treg cell-types in TCR features. Considering that Treg is a distinct T cell-type developed through a unique thymic selection process, TCR features may provide more reliable information regarding T cell identity.

To further validate our cell-type TCR signatures, we independently estimated them using the public single cell RNA-seq dataset from the COMBAT consortium data. We detected distinct clusters of T cell-types (**Figure 4a, 4b**), and estimated cell-type-specific TCR signatures using a pseudo-bulk approach (**Methods**). Although ImmuNexUT and COMBAT consortium data were generated using completely different experimental and analytical pipelines, both cell-type and TCR signatures were substantially correlated with each other (**Figure 4c**). These findings further confirmed the robustness of our signatures.

**Figure 4.**
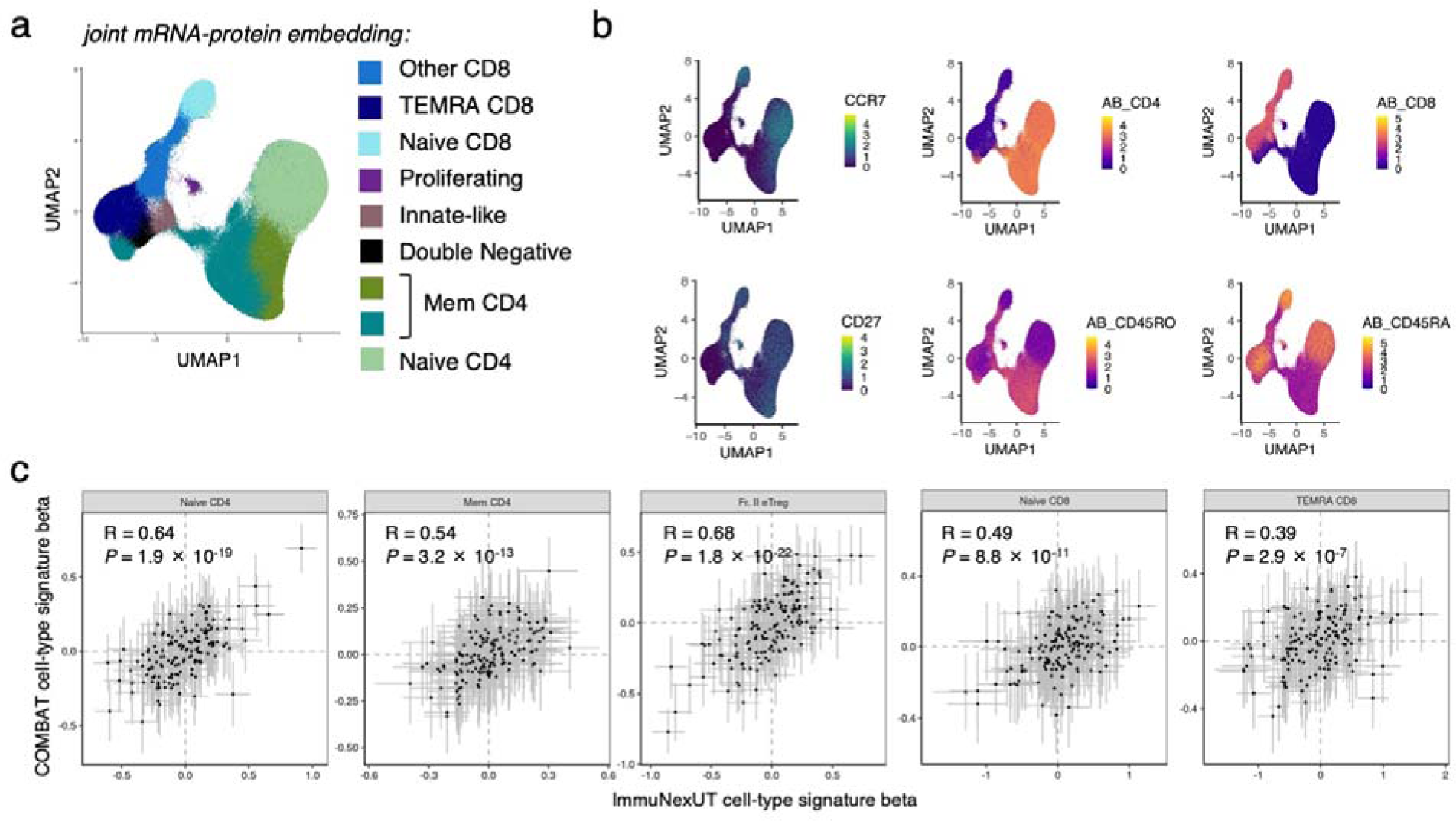
**Validation and reproducibility of TCR cell-type signature**. (a) Joint mRNA-protein embedding of COMBAT scRNA-seq data. (b) Representative feature plots of selected mRNA (CCR7 and CD27) and protein markers. (c) Comparison of the ImmuNexUT and COMBAT cell-type signatures. The correlations were evaluated with Pearson’s correlation tests.

### Disease signatures separating IMDs and HC in CD4+ T cells

We continued a supervised approach to infer ‘disease signatures’ based on transcriptome and TCR features (**Figure 1c**, **Methods**). In this analysis, we applied a linear model to compare each IMD with HCs within each T cell-types: e.g., we compared SLE Th1 with HC Th1 to obtain SLE Th1 disease signature. We controlled the effects of age, sex, and study batch (**Methods**). Importantly, *HLA* alleles can be a strong confounding factor in TCR analysis since they correlate with both IMDs’ susceptibility and TCR repertoire sequences ^26, 27^ (**Supplementary Note**). Therefore, we additionally included HLA alleles as covariates.

In the transcriptome analysis for SLE, we detected on average of 7,022 genes with differential expression, constituting 52 % of the tested genes at FDR < 0.05 (n= 13,483 genes x 13 cell-type, 175,279 tests in total, **Figure 5a**, **Supplementary** Figure 6a, and **Supplementary table 5**). The transcriptome signatures of SLE were replicated very well in the independent replication dataset (**Supplementary** Figure 3f-g). SLE Th1 showed most pronounced dysregulation in the transcriptome (**Figure 5a**). Variance partitioning analysis confirmed that the disease condition had the strongest impact on Th1 (**Supplementary Note**). For example, the expression of interferon signaling genes, particularly *IFI27* and *RSAD2*, exhibited the most substantial upregulation in SLE Th1 (**Figure 5b**). Additionally, Th1 data showed a noteworthy increase in *FOXP3* expression within the top one percentile of the disease signatures (**Figure 5b, Supplemenary table 5**). To explore the dysregulated biological processes in SLE T cells, we conducted GSEA. As expected, this analysis revealed upregulated interferon signaling pathways in many cell-types (**Figure 5c**, **Supplementary** Figure 7). In addition, multiple other pathways showed dysregulation in SLE: e.g., oxidative phosphorylation and MTORC1 signaling genes, which were pathways upregulated in regulatory T cells (**Figure 3k**), exhibited the strongest enrichment in SLE Th1 cells.

**Figure 5.**
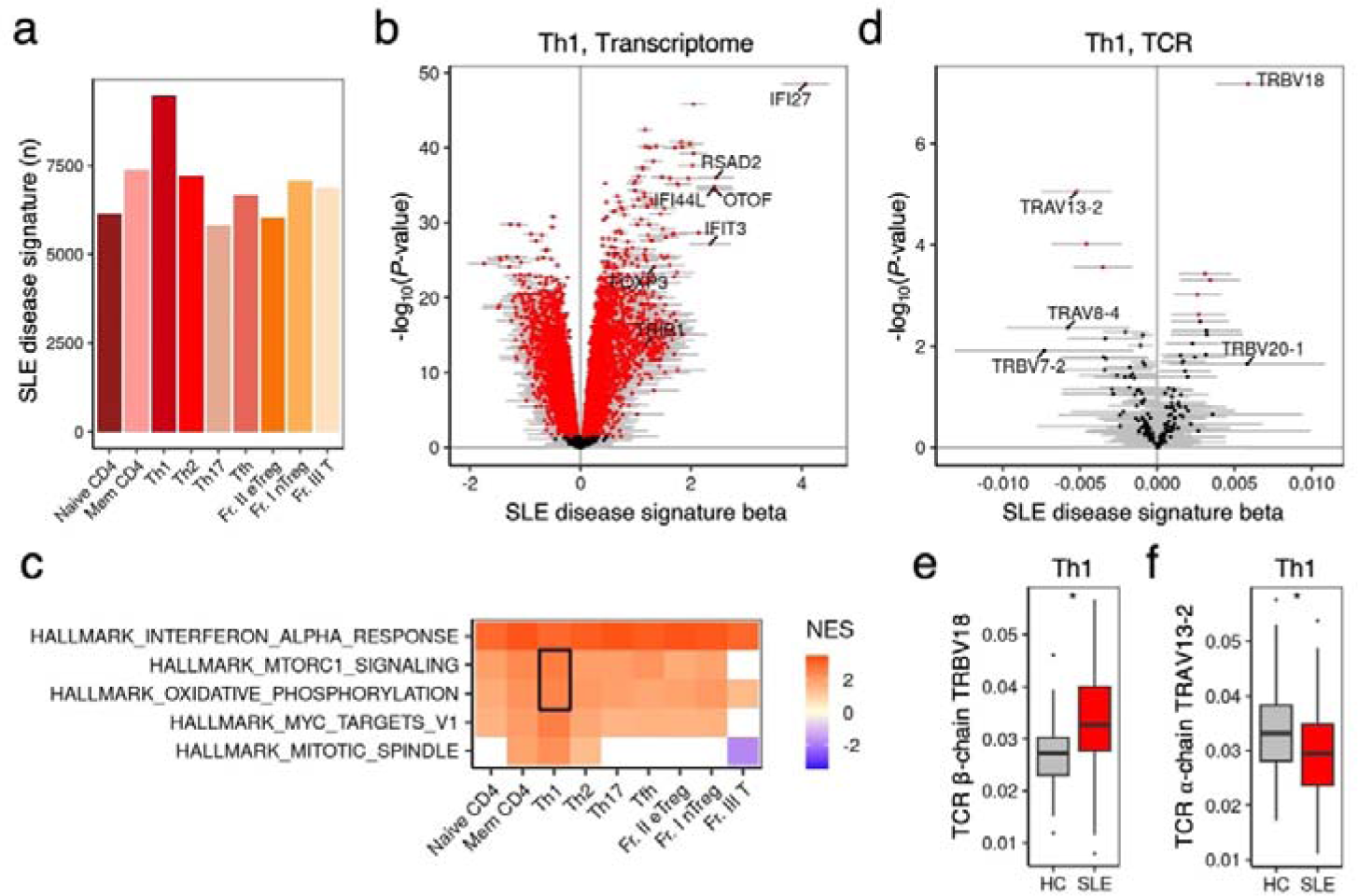
SLE disease signatures. (a) The number of significant transcriptomic SLE disease signatures in CD4+ T cell-types (at FDR < 0.05). (b,d) The disease signature of SLE Th1 by transcriptome analysis (b) and TCR analysis (d). Genes with FDR < 0.05 were noted with red color. (c) A heatmap showing normalized enrichment score (NES) of selected pathways from SLE disease signature Gene Set Enrich Analysis. (e-f) Representative boxplots showing the TCR gene usage of TRBV18 and TRAV13-2 in Th1. * indicates FDR < 0.05 in the SLE disease signature analysis.

In the TCR analysis for SLE, we detected on average 10.5 TCR features with differential usage for SLE, representing 6.6 % of the tested features at FDR < 0.05 (n= 159 feature x 13 cell-type, 2,067 tests in total, **Figure 5d**, **Supplementary** Figure 6a, **Supplementary table 6**). Although this analysis had only moderate statistical power, the results replicated well in an independent replication dataset (**Supplementary** Figure 3f**, h**). The TCR repertoire of SLE Th1 cells demonstrated increased use of TRBV18 and decreased use of TRAV13-2 (**Figure 5d-f**). For other IMDs, the number of differentially expressed TCR features was notably limited, and hence we restricted our analysis to SLE disease signatures for the downstream analyses (**Supplementary** Figure 6).

### Correlation analysis between cell-type and disease signatures

To investigate T cell plasticity in SLE, we tested correlations between CD4+ T cells’ cell-type and disease signatures. The rationale behind our strategy is that when T cell plasticity is characterized by a transition from cell-type A to cell-type B, the cell-type B disease signature should resemble the cell-type A signature. We considered both TCR and transcriptome signatures for this analysis. We found significant positive correlations between cell-type signatures of Treg and Treg-like cell-types (Fr. II eTreg and Fr. III T) and SLE disease signatures of effector CD4+ T cells (Th1, Mem CD4 and Tfh) in both TCR and transcriptome space (**Supplementary** Figure 8, **Supplementary table 7**). Among all significant correlations of Fr. II eTreg cell-type signature, SLE Th1 disease signature exhibited the largest correlation both in the TCR and transcriptome analysis (**Figure 1d**, **Figure 6a-c**).

**Figure 6.**
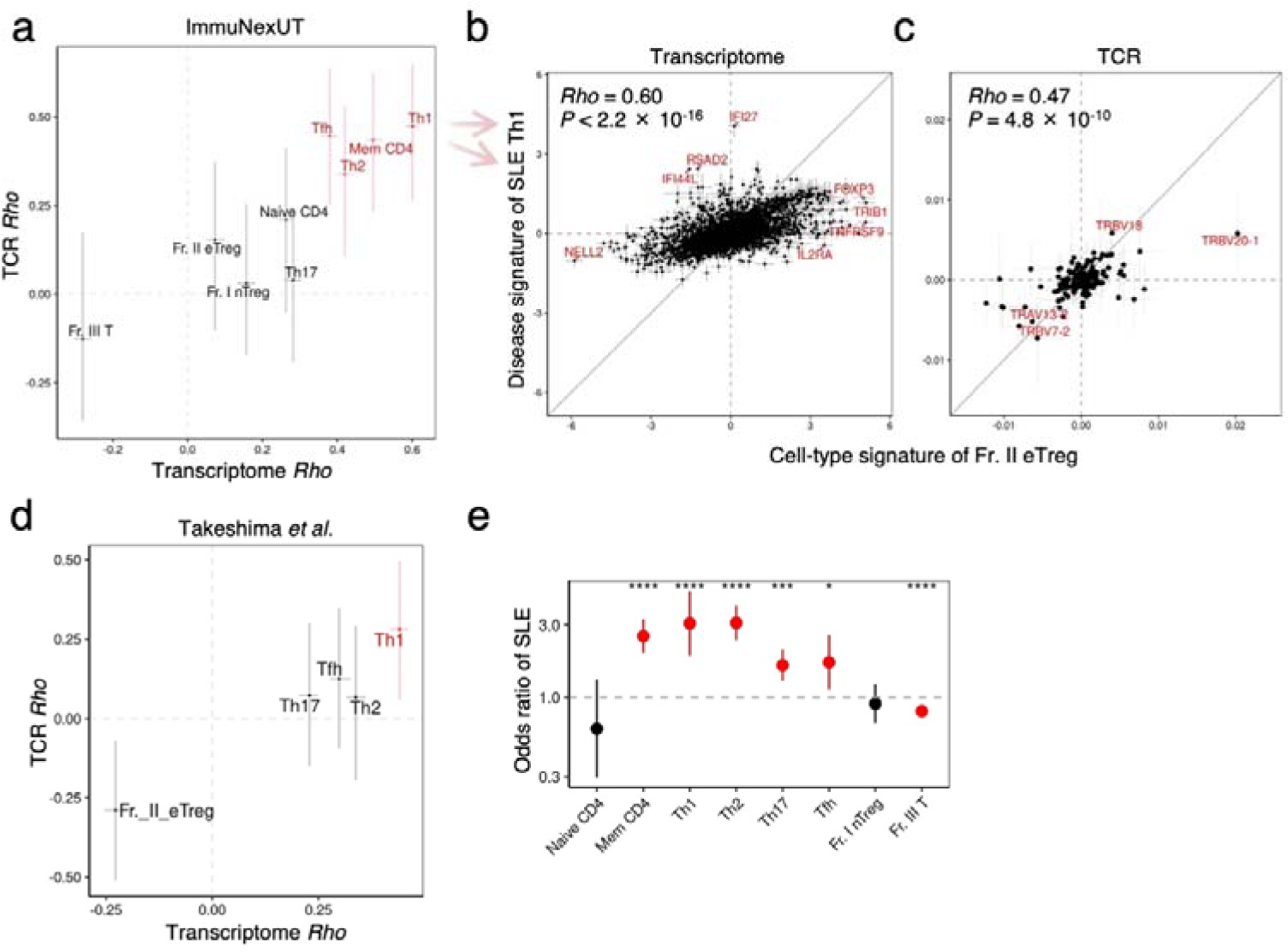
Plasticity of Fr. II eTreg to Th1 in SLE. (a) Comparison of Spearman’s correlation coefficients between Transcriptome and TCR analysis in ImmuNexUT dataset. X axis displayed the Spearman’s correlation coefficients with the 99% confidence interval, focusing on the correlation between the transcriptome Fr. II eTreg cell-type signature and SLE disease signature in CD4+ T cell-types. Y axis represented the TCR counterparts. (b-c) The correlation between Fr. II eTreg cell-type signature and SLE disease signature using transcriptome (b) and T cell receptor datasets (c) from ImmuNexUT. Grey bar in each dot indicated ±2 SE ranges. (d) Comparison of Spearman’s correlation coefficients between Transcriptome and TCR analysis in Takeshima *et al*. dataset. (e) The odds ratio of TCR clonotype overlap with Fr. II eTreg in SLE in comparison to HC in CD4+ T cell-types based on logistic mixed effect analysis. Error bars indicated 95% confidence intervals. Red color indicated P values < 0.05. * P < 0.05, *** P < 0.001, and **** P < 0.0001.

To test the replicability of our findings, we repeated the identical analysis in the replication cohort and confirmed that the strongest correlation between Fr. II eTreg cell-type signatures and SLE Th1 disease signature of both transcriptome and TCR analysis (**Figure 6d**, **Supplementary** Figure 3i-k). Importantly, we observed a much weaker correlation in the opposite direction, specifically between the Th1 cell-type signature and the SLE Fr. II eTreg disease signature, in both the TCR and transcriptome analyses (**Supplementary table 7**). Together, these results mean that effector CD4+ T cells in SLE possess T-reg like features of TCR and transcriptome and suggest a directional plasticity from regulatory T cells to effector CD4+ T cells in SLE.

### Shared TCR clonotypes between SLE cell-types

To obtain more evidence of the plasticity between CD4+ T cell-types in SLE, we quantified the TCR clonotype overlap between CD4+ T cell-types in each donor (**Methods**). We first restricted our analysis to HC samples and asked whether closely related T cell-types exhibit more TCR clonotype overlap than unrelated cell-types. As expected, we observed higher TCR clonotype overlap between Fr. II Treg and Fr. III T and between effector CD4+ T cell-types (**Supplementary** Figure 9a). This finding was consistent to our previous comparison between cell-type signatures (**Figure 3i**).

To assess the significance and magnitude of TCR clonotype overlap in SLE, we applied a generalized linear mixed model approach where we treated each TCR clonotype as an independent observation and modeled the clonotype overlap as a binary event (**Methods**). We first focused on Fr. II eTreg highlighted in the signature correlation analysis. In SLE, Fr. II eTreg showed significantly more frequent overlap with effector CD4+ T cell-types than HC (**Figure 6e**, Mem CD4, Th1, Th2 and Tfh). Among the effector cell-types, Th1 and Th2 showed the most frequent overlap: OR = 3.1, *P* = 9.1 × 10^-6^ in Th1 and OR = 3.1, *P* = 2.3 × 10^-15^ in Th2 (**Figure 6e**). Enhanced TCR overlaps between Fr. II eTreg, Th1, and Th2 were bidirectional, showing increased overlap both from Fr. II eTreg to Th1 and from Th1 to Fr. II eTreg (**Supplementary** Figure 9b). Hence, the analysis of TCR clonotype overlap also provided additional evidence for the plasticity between Fr. II eTreg and effector CD4+ T cells, including Th1.

### Clinical relevance of Th1 Tregness in SLE

The above analyses identified the Tregness of SLE Th1 T cells, possibly reflecting Fr. II eTreg-Th1 plasticity in SLE. We next sought to understand the clinical relevance of this plasticity. For this purpose, we first developed a Treg score based on the transcriptome Fr. II eTreg cell-type signature calculated by the ImmuNext HC samples (**Methods**). We applied the Tregness scoring system to the validation cohort and confirmed its robustness (**Figure 7a-b**). We then evaluated the SLE-associated Tregness of each cell-type by comparing the score of SLE and HC. Consistent with the results in Figure 6, Tregness was prominent in Th1 and other effector CD4+ T cell-types both in the ImmuNext and the replication datasets.

**Figure 7.**
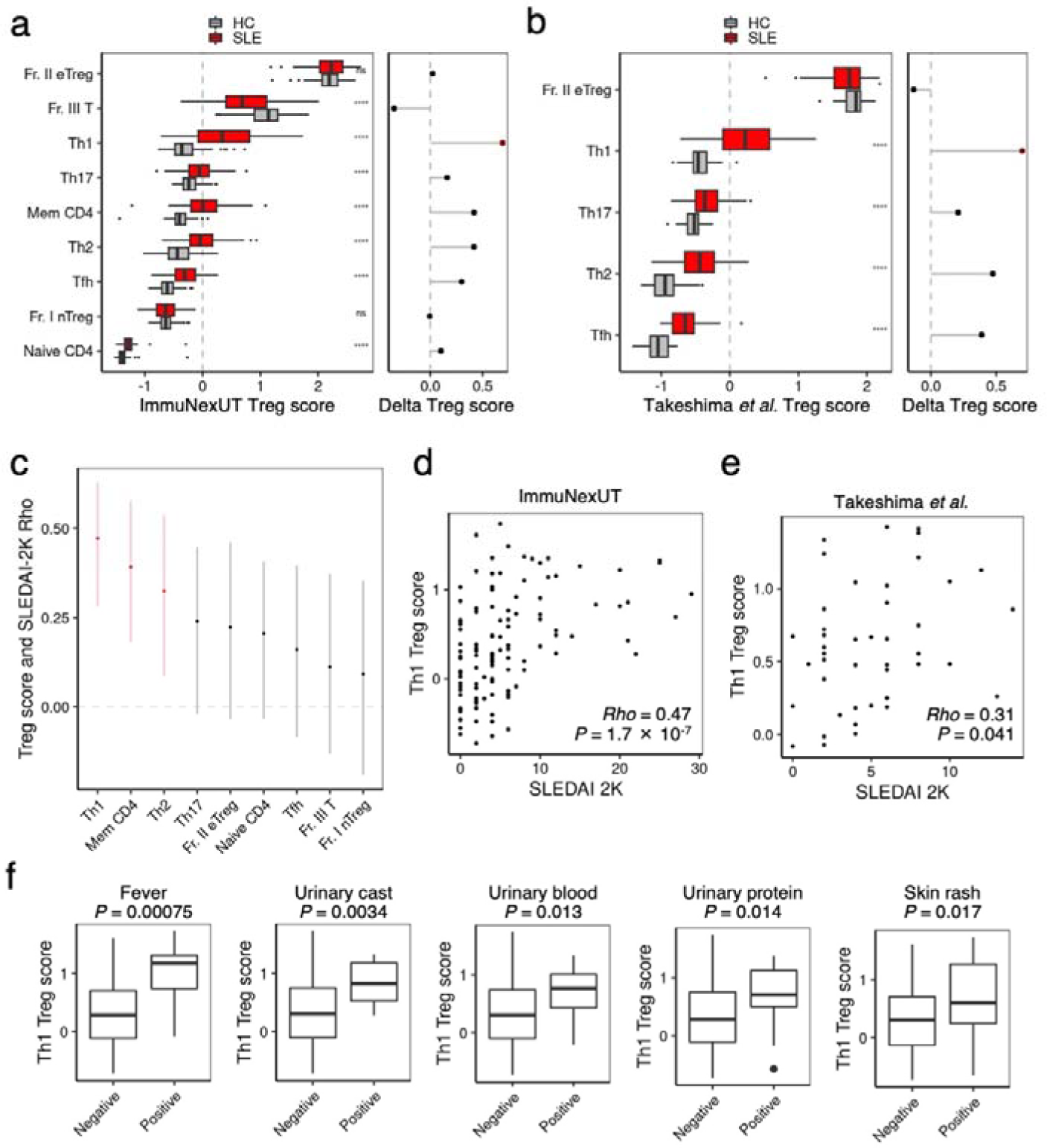
Treg score and SLE disease activity. (a-b) Treg score in CD4+ T cell-types from HC in ImmuNexUT discovery cohort (a) and Takeshima et al validation cohort (b). The difference of Treg score between HC and SLE was tested with Student’s t-tests. ns not significant, * P-value < 0.05, **** P-value < 0.0001. On the right, the delta difference in the mean Treg score between SLE and HC was shown. (c) The Spearman’s correlation coefficients with the 99% confidence intervals between Treg score in each CD4+ T cell-type and SLEDAI-2K score in the ImmuNexUT cohort. (d-e) The correlation between Treg score in Th1 and SLEDAI-2K was evaluated with Spearman’s correlation coefficients in ImmuNexUT (d) and in Takeshima et al (e). (f) Boxplots comparing the Treg scores in Th1 by the positivity of the SLEDAI-2K components in ImmuNexUT dataset. Student’s t-test was performed in the comparison of negative and positive group. Of the 24 SLEDAI-2K components, only 5 components were significantly different with P-value < 0.05.

We recently reported SLE patients’ BCR tend to be biased toward naïve B cells and proposed the scoring system to represent BCR repertoire naiveness for each sample (BCR repertoire naiveness score) ^28^. The BCR repertoire naiveness was correlated to interferon signature of SLE. To unveil the potential relationship of the interferon signature, the T cell plasticity, and B cell abnormality in SLE, we compared the Th1 interferon-α signature, the Th1 Treg score, and the BCR repertoire naiveness score. Intriguingly, we found a positive correlation between these scores (**Supplementary figure 10a-e**). This indicates a potential pathophysiological connection between the interferon signature, the T cell plasticity and BCR abnormality in the context of SLE. Additionally, the interferon signature levels in other IMDs were associated with the correlation between the Fr. II eTreg cell-type signature and the Th1 disease signature of these IMDs (**Supplementary** Figure 10f). This suggests that the interferon signature may play a role in Treg to Th1 plasticity in other IMDs as well.

Finally, we evaluated the association between Th1-Tregness and SLE clinical phenotypes. Among all CD4+ T cell-types, the Th1 Treg score exhibited the strongest association with the SLEDAI-2K score (**Figure 7c)**.

In both cohorts, the Th1 Treg score showed a positive association with the SLEDAI-2K, a composite score based on multiple clinical phenotypes, indicating a link to the global disease activity (**Figure 7d-e**). When we investigated each component of clinical phenotypes, the Th1 Treg score was significantly associated with active symptoms such as fever, nephritis, and rash (**Figure 7f**). Mediation analysis revealed that Th1-Tregness mediated the effects of the interferon signature on SLEDAI-2K (**Supplementary figure 10g-h**). These findings highlight a link between the T cell plasticity and the clinical heterogeneity in SLE and suggest the utility of the Tregness score of Th1 T cells as a SLE biomarker.

## Discussion

In this study, we thoroughly investigated the cell-type and disease signatures of purified T cell-types in SLE and other IMDs by combining TCR repertoire and transcriptome analyses. Comprehensive correlation analysis between cell-type and disease signatures provided evidence that Th1 in SLE possess Treg-like features, suggesting Treg-Th1 plasticity in SLE. TCR clonotype overlap analysis further supports the plasticity between these cell-types. We further conducted intensive replication analyses using single cell and bulk paired TCR and transcriptome data. We finally demonstrated the substantial correlation of the Th1 Tregness with clinically relevant SLE phenotypes and suggested the clinical utility of our finding as a biomarker of SLE in the clinical setting.

This study extends our previous work reporting TCR cell-type signature of Tregs^15^. A notable example is the unique Th2 TCR features characterized by hydrophobic and basic CDR3 amino acid compositions, implying a distinct differentiation machinery of Th2 cells compared to other effector CD4+ T cell-types. Semi-Unsupervised analysis through CCA also supported the existence of unique Treg and Th2 cell-type signatures. The TCR repertoire is formed through the random rearrangement in the thymus, preceding the encounters with cognate antigens. Therefore, the TCR sequence features we identify should play a causal role in the thymic or peripheral differentiation of T cells.

Ttranscriptome disease signatures, or differentially expressed genes, of SLE have been extensively researched, revealing overall upregulation of interferon signatures and various cell-type specific signatures of the disease ^19, 21, 29, 30, 31^. Our study uniquely adds cell-type-specific “TCR” disease signatures of SLE, which can serve as a tracking signature for T cell plasticity. In our analyses, we carefully controlled for the effects of the HLA alleles, which can be a major confounder associated with both IMD susceptibility and TCR repertoire patterns^26, 27^. Even using our stringent approach, we observed a substantial degree of TCR abnormality in SLE. Previous SLE TCR repertoire analysis reported different V and J gene usage in unfractionated whole blood samples^32, 33^. We successfully extended the investigation of SLE TCR abnormality to fine-grained T cell-types, which shed light on T cell plasticity in SLE.

To date, there has been scarce evidence showing T cell plasticity in human autoimmunity. Our results indicate that most prominent T cell plasticity in SLE occurs in the transition from Tregs to Th1 cells. Previous mouse studies have revealed the plasticity of Treg to effector T cells using genetically engineered tracking systems of FOXP3 protein expression^12, 13^. These studies showed that ex-FOXP3 effector T cells were pathogenic in autoimmune disease models. In our study, Treg cell-type TCR signatures, which are causally associated with Treg fate, served as a tracking signature of Treg cells in SLE. We demonstrated the association between Treg-like Th1 cells, BCR repertoire abnormalities, and disease activity in SLE. Our study illuminated the existence of Treg plasticity in human SLE, which could be pathogenic, aligning with previous reports of Treg plasticity in mice. Recently, cytotoxic CD4+ T cells were proposed to be the human counterpart of ex-FOXP3 Tregs^34^. Considering the similarity between Th1 and cytotoxic CD4+ T cells^35, 36^, such as high expression of T-bet and TCR clonotype overlap, the Treg to Th1 plasticity could be a universal phenomenon in the human immune system.

Our analytical approach to TCR data is unique in that we go beyond the molecular barcode approach to investigate individual components of the TCR sequence, which can be represented with continuous physicochemical values^37^. This approach is clearly different from a popular approach that treats TCR sequence information as a cellular barcode, focuses only on the small fraction of TCR clonotypes shared between the cell-types, and does not analyze the majority of the TCR repertoire^34, 38, 39^. With our approach, we were able to shed light on the directionality of T cell plasticity in SLE. In contrast, conventional approaches relying on clonotype overlap do not suggest any directionality (**Supplementary** Figure 9b).

Although we discovered multiple novel aspects of SLE T cell biology, we need to acknowledge some critical limitations in this study. First, we could not determine whether the T cell plasticity is causal in SLE pathogenesis or a result of the disease. To address this point, we need future longitudinal studies following TCR, transcriptome, and clinical path. Second, we did not conduct functional investigations to delve into the mechanisms underlying cell type signatures and the plasticity. Third, further exploration is needed in other IMDs to determine whether T cell plasticity is specific to SLE or more prevalent across other IMDs.

Our study provided a comprehensive picture of T cell-type plasticity from the combined viewpoint of TCR and transcriptome data at an unprecedented scale and sheds new light on the role of the T cell plasticity in the pathogenesis of SLE.

## Methods

### ImmuNxUT study cohorts

The study design and experimental details are described in the ImmuNexUT flagship article ^40^. The total number of study participants increased from n=416 in the ImmuNexUT first release data ^40^ to n=588 in this report. Study participants consisted of healthy controls (HC) and patients with the following 10 autoimmune diseases: systemic lupus erythematosus (SLE), systemic sclerosis (SSc), idiopathic inflammatory myopathy (IIM), rheumatoid arthritis (RA), ANCA-associated vasculitis (AAV), Behçet’s disease (BD), mixed connective tissue disease (MCTD), adult-onset Still’s disease (AOSD), Sjögren’s syndrome (SjS), or Takayasu arteritis (TAK) (**Supplementary Table 1**). They were recruited from five hospitals in Tokyo: the University of Tokyo Hospital, the Jikei University School of Medicine, St. Luke’s International Hospital, the National Center for Global Health and Medicine, and Tokyo Metropolitan Komagome Hospital with approval by the ethics committee at each site. Written informed consent was obtained from all participants. This study was performed in accordance with the latest version of the Helsinki declaration. This study was approved by the Ethics Committees of the University of Tokyo (G-10095).

### Peripheral blood sample collection and processing

Samples were collected in two phases. In both phases, peripheral blood mononuclear cells (PBMCs) were isolated by density gradient separation with Ficoll-Paque (GE Healthcare) immediately after the blood draw. Erythrocytes were lysed with potassium ammonium chloride buffer, and Fc-gamma receptor antibodies were used to prevent non-specific binding. In phase one, we sorted the PBMCs into seven CD4+ T cell-types, two CD8^+^ T cell-types using a MoFlo XDP instrument (Beckman Coulter) (**Supplementary Table 2**). Sorted cells were lysed and stored at -80℃. Total RNA was extracted using RNeasy Micro Kits (Qiagen). In phase two, we sorted the PBMCs into nine CD4+ T cell-types and four CD8^+^ T cell-types, using a BD FACSAria Fusion (BD Biosciences) (**Supplementary Table 2**). Sorted cells were lysed and stored at -80℃. Total RNA was extracted using MagMAX-96 Total RNA Isolation Kits (Thermo Fisher Scientific). We aimed to collect 5,000 cells per sample. All RNA-seq libraries were prepared using SMART-seq v4 Ultra Low Input RNA Kits (Takara Bio).

Genomic DNA was isolated from peripheral blood using a QIAmp DNA Blood Midi Kit (Qiagen) and whole genome sequencing libraries were prepared using TruSeq DNA PCR-Free Library Prep Kits (Illumina).

### RNA-sequencing (RNA-seq)

Prepared libraries were sequenced on HiSeq2500 or NovaSeq6000 (Illumina) to generate 100 or 150 base paired-end reads, respectively. For read level quality control of the sequencing data, adaptor sequences were trimmed with Cutadapt and reads containing too many low-quality bases (Phred quality score < 20 in more than 20% of the bases) were removed. Reads were aligned against the GRCh38 reference sequence using STAR ^41^ and gene expression was quantified with HTSeq ^42^. We also excluded samples with low uniquely mapped read rates or unique read counts less than 6 × 10^6^. For the sample level quality control, we calculated the correlation coefficient of the expression data between two samples belonging to the same cell-type then determined the average of the correlation coefficient (Di). Samples with a Di less than 0.9 were removed.

Memory CD8 cells, which isolated in phase one and divided into central memory and effector memory cell-types in phase two, were also removed from this analysis. We analyzed 6392 T cell-type RNA-seq data after quality control and the mean Di score was 0.967. After sample quality control, genes with low expression counts (< 10 in > 90% of samples) were removed. The gene expression data were normalized between samples with TMM by edgeR ^43^, and converted to log2(counts per million + 1).

In the principal component analysis, weighted variance partitioning analysis, and canonical correlation analysis, we used ComBat ^44^ to adjust for the batch effects of the study phases and RNA-seq run batches and utilized the 500 most variable genes.

### Human leukocyte antigen typing from whole genome sequencing data

Whole genome sequencing libraries were sequenced on Illumina’s HiSeq X sequencer with 151 bp pair-end reads. Reads were aligned to the reference human genome (GRCh38) by BWA-MEM based on the standardized best-practice method proposed by GATK (v 4.0.6.0) ^45^. Classical HLA alleles (HLA-A, -B, -C, -DQA, -DQB, and -DRB) were typed by graph-guided assembly software, Kourami ^27, 46^. Forty-five common HLA alleles with allele frequencies more than 0.05 were included in the analysis. The reported typing accuracy of Kourami is >98% ^27, 46^.

### TCR repertoire analysis

TCR sequences of RNA-seq data were aligned with the MiXCR (v 3.0.11) “analyze shotgun” command using default parameters ^47^. Our analysis focused solely on productive TCRs, encompassing both α and β chains. On average, each sample had 1044 unique β chain clonotypes, totaling 6.7 million across all samples. We focused on the sample level TCR sequence features in this study. V gene usage and J gene usage was defined by the usage frequency in all detected clones, regardless of the read counts in each clone. We computed the average frequency of CDR3 amino acid usages by considering the middle amino acid positions (IMGT P108-P112) in the CDR3 region for each sample ^15^. We conducted a collective analysis of these positions across varying CDR3 lengths: 11-17 for α chain CDR3 and 12-18 for β chain CDR3. TCR clones with excessively short or long CDR3 regions were excluded from this analysis.

Out of a total of 238 TCR repertoire sequence features involving V gene usage, J gene usage, and CDR3 amino acid usages from both α and β chains, we narrowed our focus to 159 TCR repertoire sequence features. These features had a mean usage frequency in all samples of over 0.01. CDR3 amino acids were categorized based on their physicochemical features ^15^.

In the principal component analysis, canonical correlation analysis, and weighted variance partitioning analysis we used ComBat to adjust for the batch effects of the study phases and RNA-seq run batches and utilized the 50 most variable TCR repertoire sequence features.

### Regularized canonical correlation analysis

To unlock the complex relationships among many variables in large TCR and transcriptome datasets, we performed regularized canonical correlation analysis (CCA) of the two datasets. CCA is a dimension-reduction method that quantifies the covariation between “two” datasets and accordingly project each dataset into low-dimensional space: i.e., canonical variates (CVs). We conducted regularized Canonical Correlation Analysis (CCA) using the R mixOmics package (v6.20.0), with ridge penalties tuned via 10-fold cross-validation. The paired datasets were projected to paired 10 Canonical Variables (CVs). We utilized CVs derived from the TCR dataset, unless specified otherwise.

To evaluate the canonical correlation values, we permuted the TCR and transcriptome data 100 times and empirically compared the observed canonical correlations to the values from the permutations.

In the “inter-cell-type” CCA analysis, we utilized datasets from HC samples to avoid any influences from IMD disease status (1,366 samples from 134 donors). We then conducted “intra-cell-type” CCA to ask whether TCR and transcriptome data captured the shared variation component separating IMD cases and HCs within each cell-type.

To identify cell-type-associated CVs, we tested the association of each identified inter-cell-type CV of CD4+ and CD8+ T cell-types with corresponding cell-types using a linear model:

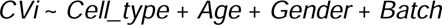

where *CVi* represents canonical variable 1 to 10 in CD4+ or CD8+ T cell-types, *Cell_type* represents the categorical value of the specific cell-type, such as Th1, in comparison to all other CD4+ or CD8+ T cell-types, *Age* is the age of the individual at the time of recruitment, *Gender* indicates categorical sex of the individual, and *Batch* represents two study phases. CVs that were associated with at least one cell-type with a threshold of FDR < 0.05 were considered cell-type-associated CVs.

To identify disease-associated CVs, we tested the association of each identified intra-cell-type CVs with IMD using a linear model:

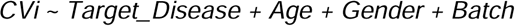

where *CVi* represents canonical variable 1 to 10 in each T cell-types, *Target_Disease* represents the categorical value of the specific IMD, such as SLE, in comparison to HC, and other variables maintain previous definitions. CVs that were associated with at least one IMD with a threshold of FDR < 0.05 were considered disease-associated CVs.

Next, we assessed the correlations between these cell-type-associated inter-cell-type CVs and disease-associated intra-cell-type CVs using Pearson’s correlation test. In the correlation analysis, we employed loading vectors of the CVs, which reflect the biological significance of each variable in the CCA projection. Because both positive and negative correlations are significant, we compared the R^2^ values of the correlations between cell-type-associated inter-cell-type CVs and disease-associated intra-cell-type CVs, and between cell-type-non-associated inter-cell-type CVs and disease-non-associated intra-cell-type CVs (background).

### Weighted variance partitioning analysis

To calculate the explained variance of each factor for whole transcriptome or TCR repertoire sequence variation of HC, we fitted the each PC score to the linear mixed models in R variancePartition package (v1.26.0) ^48^ and then inferred the average value of the explained variance weighted by each PC’s eigenvalue for each analysis ^19^.

Initially, we assessed whether the ComBat method effectively minimizes the recognized batch effects in the study phases. This assessment involved utilizing the subsequent linear mixed model formula for both the data prior to and after implementing the ComBat procedure for study phases and RNA-seq run batches.

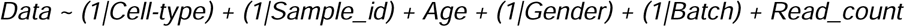

where *Data* represents either transcriptome or TCR repertoire sequence feature dataset, *Cell-type* is the categorical value of T cell-types, *Sample_id* is the categorical value assigned to each individual, *Age* is the age of the individual at the time of recruitment, *Gender* indicates categorical sex of the individual, *Batch* represents two study phases, and *Read_count* is the count of unique map reads from RNA-seq data after quality control.

After discovering that the explained variance attributed to Batch and Read_count is insignificantly low following the implementation of the ComBat procedure, we applied the subsequent formula to the datasets that underwent the ComBat process:

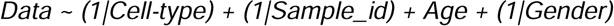

where variables maintain previous definitions.

To compute the variance explained by each factor in the variation of the transcriptome or TCR repertoire sequences within each T cell-type, we conducted analysis using samples from both HC and individuals with IMDs. We initially performed ComBat method to minimizes the recognized batch effects in the study phases and RNA-seq run batches.

Then, we fitted the each PC score to the linear mixed models in R variancePartition package (v1.26.0) and inferred the average value of the explained variance weighted by each PC’s eigenvalue for each analysis. we applied the subsequent formula to the datasets that underwent the ComBat process:

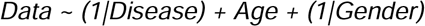

where *Disease* represents the categorical value of IMDs and other variables maintain previous definitions.

Additionally, to compare inter-cell-type and intra-cell-type heterogeneity of the TCR dataset, we modeled PC score of all TCR dataset in the following model:

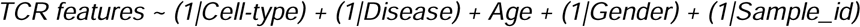

where variables maintain previous definitions. We compared the average explained variance value weighted by each PC’s eigenvalue. In this analysis, we considered that the explained variance by *Cell-type* represents the inter-cell-type heterogeneity, and the explained variance by *Disease* represents the intra-cell-type heterogeneity of the dataset.

In the weighted variance partitioning analysis of the CCA, we fitted the each TCR CVs to the identical linear mixed models of the transcriptome or TCR feature analysis. We calculated the total amount of variance that can be explained by CV_i_:

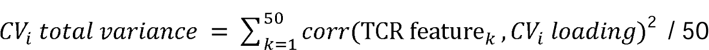

Where *TCR feature_k_* represents the vector of one of the scaled TCR features and *CVi loading* represents the TCR loading vector of CV_i_. Then, we inferred the average value of the explained variance weighted by each CV’s total variance.

To verify whether the inferred explained variance was not biased by outlier individuals, we estimated the distribution of the explained variance by jackknife resampling method. When we had *n* individuals for one analysis, we re-calculated the explained variance *n* times by excluding all samples from each individual and estimated the 95% confidence intervals of the explained variance.

### CD8+ T cells signature analysis

To understand the characteristics of CD8+ T cell signatures compared to CD4+ T cells in HC, we employed two analyses. The first analysis involved a standard linear model approach:

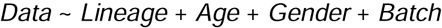

where *Data* represents either transcriptome or TCR repertoire seqeuence feature dataset, *Lineage* is the categorical value of CD8+ or CD4+ T cells, *Age* is the age of the individual at the time of recruitment, *Gender* indicates categorical sex of the individual, and *Batch* represents two study phases.

The second was linear mixed model analyses conditioned the effects of donor batches and T cell-type batches calculated by lmer function of lme4 package:

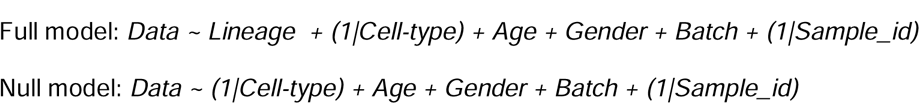

where *Cell-type* is the categorical value of T cell-types, *Sample_id* is the categorical value assigned to each individual, and other variables maintain previous definitions.

We used the beta and standard error (SE) values of the full model. *P*-values were calculated through an analysis of variance performed on both the full and null models.

### Cell-type signature analysis

We performed a linear mixed model analysis, conditioning the effects of age, sex, and donor batch effects, using the CD4+ or CD8+ T cells samples from HCs:

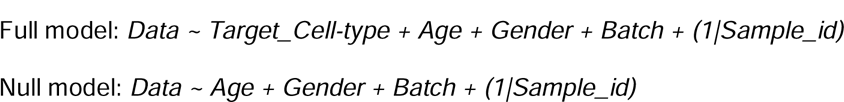

where *Data* represents either transcriptome or TCR repertoire sequence feature dataset, “*Target_Cell-type*” represents the categorical value of the specific T cell-type, such as Fr. II eTreg, in comparison to other CD4+ T cell-types, *Age* is the age of the individual at the time of recruitment, *Gender* indicates categorical sex of the individual, and *Batch* represents two study phases, and *Sample_id* is the categorical value assigned to each individual.

We used the beta and SE values of the full model. *P*-values were calculated through an analysis of variance performed on both the full and null models. The results were compared to simple linear model below:

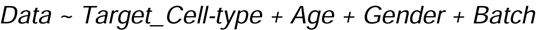

where variables maintain previous definitions.

In order to affirm the validity of the linear mixed model investigations concerning the association between *Target_Cell-type* and *Data*, a series of 1000 permutation tests were conducted. During these tests, the T cell-types within each individual were randomly rearranged, maintaining the existing data configuration of the other explanatory factors.

### Analysis of associations between HLA alleles and TCR repertoire sequence features

We conducted an association analysis between HLA alleles and TCR repertoire sequence features using only HC samples, following the analysis pipeline we previously described ^27^. In each T cell-types, a linear model analysis accounting for the effects of HLA allele dose (0-2), age, gender, and study phase batch were conducted.

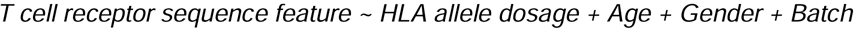

where *T cell receptor sequence feature* is the TCR repertoire sequence feature dataset, *HLA allele dosage* is the numerical value of HLA allele dosages [0-2], *Age* is the age of the individual at the time of recruitment, *Gender* indicates categorical sex of the individual, and *Batch* represents two study phases.

Statistical significance was set at FDR < 0.05 with the Benjamini-Hochberg method. In the subsequent TCR disease signature analyses, we have made adjustments for all HLA alleles that displayed significant associations with TCR sequence features in at least one T cell-type.

### Disease signature analysis

In case of transcriptome analysis, we performed a linear model analysis, conditioning the effects of age, sex, and study batch effects, using the CD4+ or CD8+ T cells cell-type samples from HCs and the target disease:

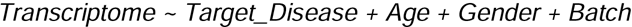

where *Transcriptome* represents transcriptome dataset, *Target_Disease* represents the categorical value of the specific IMD, such as SLE, in comparison to HC, *Age* is the age of the individual at the time of recruitment, *Gender* indicates categorical sex of the individual, and *Batch* represents two study phases.

In case of TCR analysis, we performed a linear model analysis, conditioning the effects of age, sex, study batch effects, and specific HLA alleles using the CD4+ or CD8+ T cells cell-type samples from HCs and the target disease:

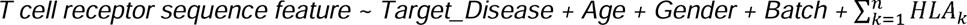

where *T cell receptor sequence feature* represents the TCR repertoire sequence feature, *Target_Disease* represents the categorical value of the specific IMD, such as SLE, in comparison to HC, *Age* is the age of the individual at the time of recruitment, *Gender* indicates categorical sex of the individual, *Batch* represents two study phases, *n* represents the number of TCR repertoire sequence feature associated HLA alleles in any of the T cell-types, and *HLA_k_* represents HLA allele in association with the TCR repertoire sequence feature.

If we have longitudinal samples, we only used samples at baseline in this analysis.

### Gene set enrichment analysis

Gene Set Enrichment Analysis (GSEA) was performed for the characterization of transcriptomic cell-type and disease signatures, usnig the MSigDB hallmark gene set (msigdbr v7.4.1) ^49, 50^. We used the beta values for the input.

### Correlation analysis between cell-type and disease signatures

Correlation analysis was conducted using nonparametric Spearman’s correlation to assess the association with either transcriptome or TCR signatures. We did not conduct this analysis in CD8+ T cell-types because, in 4 of the 10 IMDs, the study included only Naïve CD8 T cells, and the disease signatures for CD8+ T cells were restricted to Naïve CD8 T cells. We calculated the confidence interval of the correlation coefficient (Rho) through 1000 bootstraps using the rcompanion package.

### TCR clonotype overlap analysis

We analyzed individual TCR β chain clonotypes and compared them between CD4+ T cell-types in each individual. TCR clonotype overlap was defined by the overlap ratio in all detected clones, regardless of the read counts in each clone. The relationship between single clonotype overlaps and SLE disease was modeled using GLMM to control the effects of donor batches:

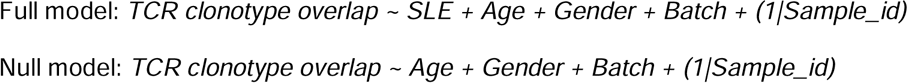

where *TCR clonotype overlap* is the categorical value of the individual TCR clonotype overlap, *SLE* is the categorical value assigned to SLE in contrast to HC, and other variables maintain previous definitions. *P*-values were calculated through an analysis of variance performed on both the full and null models. Each T cell-type was individually modeled to identify the T cell-type in which SLE case-control status significantly improves the model fit.

### Calculation of the Treg score in Th1

We utilized transcriptome data from the Th1 cell-type for scoring. Additionally, we utilized the effect estimates of the cell-type Fr. II eTreg signature at the threshold of FDR 0.05 (12088 genes). The Treg score was calculated using the following formula.

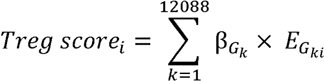

where β*G_k_* corresponded to the effect size of the *G_k_* gene in the Fr. II eTreg signature, and *EG_ki_* corresponded to the log2 transformed expression of the *G_k_* gene in individual i. Treg score was mean-centered and variance-standardized before comparison with other data.

### Interferon-α signature and B cell receptor repertoire naiveness score

The Th1 interferon-α signature score was calculated usings Gene Set Enrichment Analysis (GSEA) ^51^. We utilized the genes from HALLMARK_INTERFERON_ALPHA_RESPONSE of the MSigDB (msigdbr v7.4.1) ^49^. The B cell receptor repertoire naiveness score was calculated based on the VDJ gene usage in the B cell receptor repertoire, as we recently reported ^28^. Using the SLE patients’ data, the relationships between Th1 interferon-α signature, Th1 Treg score, and SLEDAI-2K were tested by mediation analysis using R mediation package (v4.5.0). Interferon-α signature was treated as the independent variable, Th1 Treg score as the mediator, and the SLEDAI-2K score as the dependent variable. P values were calculated via 1000-time bootstrapping.

### Takeshima et al. validation cohort analysis

The public CD4+ T cell-type RNA-seq data from 37 HC and 49 SLE patients ^21^ were reanalyzed using the identical pipeline described above. In brief, we leveraged publicly available RNA-seq data (NBDC hum0214, dataset: JGAD000371) and reanalyzed TCR repertoire sequence features in each sample. Cell-type and disease gene and TCR signatures were independently analyzed and then compared with our original discovery ImmuNexUT cohort. Naïve and memory CD4+ T cell-types were excluded from the analysis due to a different gating strategy in flow cytometric purification. Th1 Treg score was computed similarly, utilizing the Fr. II eTreg signature from the ImmuNexUT cohort.

### COMBAT dataset analysis

We identified CD4+ Tregs, CD4+ naive T cells, CD4+ memory T cells, CD8+ naive T cells, CD8+ effector T cells, and other CD8+ T cells in the COMBAT dataset. To identify CD4 Tregs, we applied unimodal dimensionality reduction: following log10CPK normalization and scaling of mRNA counts, we applied PCA (R package “irlba,” v2.3.3) to the union of the 200 most variable genes in each sample, used Harmony (v1.0) to correct PCA embeddings for batch effects, built a shared nearest neighbor (SNN) graph (R package “singlecellmethods”, v0.1.0) based on these batch-corrected embeddings, and then applied modularity clustering to this SNN graph (R package “Seurat” v3.2.2, resolution 0.5). One cluster was distinct in high *FOXP3*, *CTLA4* and *CD4* expression; we annotated this cluster as CD4+ Tregs. To identify the other cell types, we applied a multimodal dimensionality reduction pipeline^52^, incorporating both mRNA and surface protein information. The multimodal pipeline followed the unimodal pipeline described above, with CLR-normalization of protein counts and CCA instead of PCA. CCA (R package “CCA” v1.2.1) was applied to the paired mRNA and protein measurements per cell (4423 variable genes and 10 surface proteins, respectively). To emphasize traditional T cell populations, we selected 10 surface proteins critical to distinguish between CD4, CD8, central memory (CM) and effector memory (EM) T cells (AB_CD4, AB_CD8, AB_CD45RO, AB_CD45RA, AB_CD197 _CCR7, AB_CD62L, AB_CD27, AB_CD11a, AB_CX3CR1, AB_KLRG1_MAFA). We then used Harmony to remove batch effects from the mRNA-based canonical variate (CV) scores, constructed a SNN graph based on the 10 batch-corrected CVs, and applied modularity clustering as above to identify cell type clusters.

We then used a pseudo-bulk approach to estimate TCR signatures. We analyzed CD4+ and CD8+ T cells separately, following the same analysis procedure: 1) we combined all cells of the same cell type in each individual to calculate the frequency of each TCR feature, and 2) for each cell type of interest, we estimated the following mixed effects linear regression:

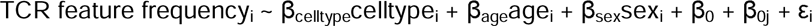

Each observation i represents one T cell type from one individual. celltype_i_ equals 1 if cell i is observed in the T cell state of interest; 0 otherwise. β_0_ is the global intercept; β_0j_ is a modification to the global intercept fit for each individual j, and ε_i_ is the error term.

## Acknowledgements

We gratefully acknowledge the invaluable support of the ImmuNexUT consortium members for their contributions to sample and clinical data collection. The super-computing resource was provided by Human Genome Center, Institute of Medical Sciences, The University of Tokyo (http://sc.hgc.jp/shirokane.html).

## Author Contributions

KI and KF designed the study. YN conducted bioinformatics analysis with the help of MN, KL, MO, HH, HT, TI, HI, SR, and KI. KL and SR contributed to COMBAT data analysis and provided critical review and revisions to the manuscript. TO, KF, and KI managed the project. YN wrote the first manuscript with critical inputs from MN, KL, HH, HT, SR, TO, KF, and KI. All authors contributed to the final version of the manuscript and approved it.

## Data availability statement

The cell-type and SLE disease signature data are provided as supplementary tables. The datasets generated during this study are available upon acceptance at the National Bioscience Database Center (NBDC) with the study accession code of hum0214 and the dataset code of JGAD000903.

## Code availability statement

We used publicly available software for the analyses. Custom codes are available upon acceptance at https://github.com/YNagafuchi/ImmuNexUT_TCR.

## References

1. Lisnevskaia, L., Murphy, G. & Isenberg, D. Systemic lupus erythematosus. Lancet 384, 1878–1888 (2014).

2. He, J. et al. Circulating precursor CCR7(lo)PD-1(hi) CXCR5⁺ CD4⁺ T cells indicate Tfh cell activity and promote antibody responses upon antigen reexposure. Immunity 39, 770–781 (2013).

3. 3. Bocharnikov, A.V., et al. PD-1hiCXCR5- T peripheral helper cells promote B cell responses in lupus via MAF and IL-21. JCI Insight 4 (2019).

4. Arazi, A. et al. The immune cell landscape in kidneys of patients with lupus nephritis. Nat Immunol 20, 902–914 (2019).

5. 5. Dunlap, G.S., et al. Single-cell transcriptomics reveals distinct effector profiles of infiltrating T cells in lupus skin and kidney. JCI Insight 7 (2022).

6. Miyara, M. et al. Functional delineation and differentiation dynamics of human CD4+ T cells expressing the FoxP3 transcription factor. Immunity 30, 899–911 (2009).

7. Yang, J. et al. Th17 and natural Treg cell population dynamics in systemic lupus erythematosus. Arthritis Rheum 60, 1472–1483 (2009).

8. Ma, X. et al. Expansion of T follicular helper-T helper 1 like cells through epigenetic regulation by signal transducer and activator of transcription factors. Ann Rheum Dis 77, 1354–1361 (2018).

9. He, J. et al. Low-dose interleukin-2 treatment selectively modulates CD4(+) T cell subsets in patients with systemic lupus erythematosus. Nat Med 22, 991–993 (2016).

10. Li, H., Boulougoura, A., Endo, Y. & Tsokos, G.C. Abnormalities of T cells in systemic lupus erythematosus: new insights in pathogenesis and therapeutic strategies. J Autoimmun 132, 102870 (2022).

11. Maecker, H.T., McCoy, J.P. & Nussenblatt, R. Standardizing immunophenotyping for the Human Immunology Project. Nature reviews.Immunology 12, 191–200 (2012).

12. Komatsu, N. et al. Pathogenic conversion of Foxp3+ T cells into TH17 cells in autoimmune arthritis. Nature medicine 20, 62–68 (2014).

13. Zhou, X. et al. Instability of the transcription factor Foxp3 leads to the generation of pathogenic memory T cells in vivo. Nat Immunol 10, 1000–1007 (2009).

14. Dominguez-Villar, M., Baecher-Allan, C.M. & Hafler, D.A. Identification of T helper type 1-like, Foxp3+ regulatory T cells in human autoimmune disease. Nat Med 17, 673-675 (2011).

15. Lagattuta, K.A. et al. Repertoire analyses reveal T cell antigen receptor sequence features that influence T cell fate. Nat Immunol 23, 446–457 (2022).

16. Li, H.M. et al. TCRβ repertoire of CD4+ and CD8+ T cells is distinct in richness, distribution, and CDR3 amino acid composition. J Leukoc Biol 99, 505–513 (2016).

17. Emerson, R. et al. Estimating the ratio of CD4+ to CD8+ T cells using high-throughput sequence data. J Immunol Methods 391, 14–21 (2013).

18. Ota, M. et al. Dynamic landscape of immune cell-specific gene regulation in immune-mediated diseases. Cell (2021).

19. Nakano, M. et al. Distinct transcriptome architectures underlying lupus establishment and exacerbation. Cell 185, 3375–3389.e3321 (2022).

20. Lagattuta, K.A., Nathan, A., Rumker, L., Birnbaum, M.E. & Raychaudhuri, S. The T cell receptor sequence influences the likelihood of T cell memory formation. bioRxiv (2023).

21. Takeshima, Y. et al. Immune cell multi-omics analysis reveals contribution of oxidative phosphorylation to B cell functions and organ damage of lupus. bioRxiv, 2021.2010.2008.463629 (2021).

22. Ferraro, A. et al. Interindividual variation in human T regulatory cells. Proc Natl Acad Sci U S A 111, E1111–1120 (2014).

23. Radens, C.M., Blake, D., Jewell, P., Barash, Y. & Lynch, K.W. Meta-analysis of transcriptomic variation in T-cell populations reveals both variable and consistent signatures of gene expression and splicing. RNA 26, 1320–1333 (2020).

24. Zeng, H. et al. mTORC1 couples immune signals and metabolic programming to establish T(reg)-cell function. Nature 499, 485–490 (2013).

25. Kempkes, R.W.M., Joosten, I., Koenen, H.J.P.M. & He, X. Metabolic Pathways Involved in Regulatory T Cell Functionality. Front Immunol 10, 2839 (2019).

26. Ishigaki, K. et al. HLA autoimmune risk alleles restrict the hypervariable region of T cell receptors. Nat Genet 54, 393–402 (2022).

27. Nagafuchi, Y. et al. Control of naive and effector CD4 T cell receptor repertoires by rheumatoid-arthritis-risk HLA alleles. J Autoimmun 133, 102907 (2022).

28. Ota, M. et al. Multimodal repertoire analysis unveils B cell biology in immune-mediated diseases. Ann Rheum Dis 82, 1455–1463 (2023).

29. Bennett, L. et al. Interferon and granulopoiesis signatures in systemic lupus erythematosus blood. J Exp Med 197, 711–723 (2003).

30. Baechler, E.C. et al. Interferon-inducible gene expression signature in peripheral blood cells of patients with severe lupus. Proc Natl Acad Sci U S A 100, 2610–2615 (2003).

31. Lyons, P.A. et al. Novel expression signatures identified by transcriptional analysis of separated leucocyte subsets in systemic lupus erythematosus and vasculitis. Ann Rheum Dis 69, 1208–1213 (2010).

32. Liu, X. et al. T cell receptor beta repertoires as novel diagnostic markers for systemic lupus erythematosus and rheumatoid arthritis. Ann Rheum Dis 78, 1070–1078 (2019).

33. Hou, X. et al. Characterisation of T and B cell receptor repertoire in patients with systemic lupus erythematosus. Clin Exp Rheumatol 41, 2216–2223 (2023).

34. Freuchet, A. et al. Identification of human exT. Nat Immunol 24, 1748–1761 (2023).

35. Cenerenti, M., Saillard, M., Romero, P. & Jandus, C. The Era of Cytotoxic CD4 T Cells. Front Immunol 13, 867189 (2022).

36. Goto, M. et al. Age-associated CD4. Sci Immunol 9, eadk1643 (2024).

37. Pauken, K.E. et al. TCR-sequencing in cancer and autoimmunity: barcodes and beyond. Trends Immunol 43, 180–194 (2022).

38. Bilate, A.M. et al. T Cell Receptor Is Required for Differentiation, but Not Maintenance, of Intestinal CD4. Immunity 53, 1001–1014.e1020 (2020).

39. Abbas, H.A. et al. Single cell T cell landscape and T cell receptor repertoire profiling of AML in context of PD-1 blockade therapy. Nat Commun 12, 6071 (2021).

40. Ota, M. et al. Dynamic landscape of immune cell-specific gene regulation in immune-mediated diseases. Cell 184, 3006–3021.e3017 (2021).

41. Dobin, A. et al. STAR: ultrafast universal RNA-seq aligner. Bioinformatics (Oxford, England) 29, 15–21 (2013).

42. Anders, S., Pyl, P.T. & Huber, W. HTSeq--a Python framework to work with high-throughput sequencing data. Bioinformatics (Oxford, England) 31, 166–169 (2015).

43. Robinson, M.D., McCarthy, D.J. & Smyth, G.K. edgeR: a Bioconductor package for differential expression analysis of digital gene expression data. Bioinformatics (Oxford, England) 26, 139–140 (2010).

44. Leek, J.T., Johnson, W.E., Parker, H.S., Jaffe, A.E. & Storey, J.D. The sva package for removing batch effects and other unwanted variation in high-throughput experiments. Bioinformatics (Oxford, England) 28, 882–883 (2012).

45. DePristo, M.A. et al. A framework for variation discovery and genotyping using next-generation DNA sequencing data. Nat Genet 43, 491–498 (2011).

46. Lee, H. & Kingsford, C. Kourami: graph-guided assembly for novel human leukocyte antigen allele discovery. Genome Biol 19, 16 (2018).

47. Bolotin, D.A. et al. MiXCR: software for comprehensive adaptive immunity profiling. Nat Methods 12, 380–381 (2015).

48. Hoffman, G.E. & Schadt, E.E. variancePartition: interpreting drivers of variation in complex gene expression studies. BMC Bioinformatics 17, 483 (2016).

49. Liberzon, A. et al. The Molecular Signatures Database (MSigDB) hallmark gene set collection. Cell Syst 1, 417–425 (2015).

50. Wu, T. et al. clusterProfiler 4.0: A universal enrichment tool for interpreting omics data. Innovation (N Y) 2, 100141 (2021).

51. Hänzelmann, S., Castelo, R. & Guinney, J. GSVA: gene set variation analysis for microarray and RNA-seq data. BMC Bioinformatics 14, 7 (2013).

52. Nathan, A. et al. Multimodally profiling memory T cells from a tuberculosis cohort identifies cell state associations with demographics, environment and disease. Nat Immunol 22, 781–793 (2021).

