## Supplementary Note for "T cell plasticity in systemic lupus erythematosus revealed by large-scale T cell receptor repertoire and transcriptome studies"

**T cell receptor and Transcriptome datasets**

Following quality control analysis using transcriptome data alone (**Methods**), 6,392 samples from 568 donors remained in total. By MiXCR T cell receptor (TCR) alignment, we found 876 and 1,034 distinct functional clonotypes in the mean from alpha and beta chains, respectively. Out of a total of 238 TCR features involving V gene usage, J gene usage, and complementary determining region 3 (CDR3) amino acid usages from both α and β chains, we extracted 159 TCR repertoire sequence features that had a mean usage frequency in all samples of over 0.01: 33 CDR3 middle position amino acid frequencies, and 69 V and 57 J gene usage frequencies. The average length of CDR3 in α-chains was 13.6, which is slightly shorter than the average length of 14.4 in β chains, and the criteria for CDR3 length were applied differently for amino acid usage analyses: 11-17 for α chain CDR3 and 12-18 for β chain CDR3 (**Extended Figure 1**).

To minimize the batch effects of the study phases and RNA-seq run batches, we applied ComBat procedure to TCR and transcriptome datasets. The ComBat procedure successfully minimized the impact of known batch effects from the study phases, and any influence from RNA-seq read count variation was found to be negligible (**Extended Figure 2**).

**Principal component analysis of transcriptome and TCR datasets**

To overview the multi-dimentional data of the transcriptome and TCR features, we first performed principal component analysis (PCA) (**Supplementary Figure 1**). In this analysis, we exclusively used samples from HC to prevent any influence from immune-mediated disease (IMDs, sample n=1,366 from 134 individuals).

By utilizing only the transcriptome data, we were able to differentiate between individual CD4+ and CD8+ T cell-types, although the distinction between CD4+ and CD8+ T cells was relatively small (**Supplementary Figure 1b**). When we performed PCA using only CD4+ T cell samples, effector CD4+ T cell-types, that are Th1, Th2, Th17, Tfh and Mem CD4, were similar to each other (**Supplementary Figure 1c**). The four CD8+ T cell-types were distinguished by PC1 and PC2, which corresponded to effectorness and memoryness respectively (**Supplementary Figure 1d**).

When we summarize the TCR features from HC samples through PCA, a significant distinction between CD4+ and CD8+ T cells emerged, which was distinct from the findings of the transcriptome analysis (**Supplementary Figure 1b and 1e**). PCA of CD4+ T cells exhibited a distinction between regulatory CD4+ T cell-types (Fr. II eTreg, Fr. I nTreg, and Fr. III T) and the remaining cell-types (**Supplementary Figure 1f)**. The TCR features of Th2 exhibited certain similarities to Tregs, in consistent to cell-type signature analysis (**Figure 2**). In contrast, the differentiation between CD8+ T cell-types was less distinct in the TCR features compared to the transcriptome (**Supplementary Figure 1d and 1g**). Thus, deciphering the TCR sequence feature information provided us with additional and complementary insights into T cells, including the distinctive TCR sequence features of CD4+ and CD8+ T cells.

**Inter-cell-type weighted variance partitioning analysis of transcriptome and TCR datasets**

We further evaluated how the transcriptome and TCR feature variations reflected the heterogeneity of the samples. We employed weighted variance partitioning analysis to quantify the impact of T cell-types and individual variations on both the transcriptome and TCR datasets (**Methods**).

Different T cell lineages (either CD4+ or CD8+) explained 20.7 % of the most variable 500 genes of the transcriptome and they explained 27.5 % of the variation of most variable 50 TCR sequence features (**Supplementary Figure 1h**), revealing the strong impact of the T cell lineage on the TCR feature variation.

Different T cell-types explained 77.5% of the transcriptome variation and they explained 39.4% of the TCR feature variation when analyzing CD4+ and CD8+ T cells together (“All” of **Supplementary Figure 1i**). Interestingly, donor batch (Sample_id) explained 15.4 % of the TCR repertoire variation in comparison 6.7 % of the transcriptome data, reflecting the strong donor batch impact on TCR features. The gender explained 2.7 % variation of the transcriptome, while it affected only the 0.082 % variance of the TCR features. These findings emphasize the added value of TCR repertoire analysis for characterizing T cell-types and assessing individual variations.

**CD8+ T cell lineage signature analysis**

Inspired by the distinct TCR features contrast between CD4+ and CD8 T+ cells (**Supplementary Figure 1e and 1h**), we systematically examined CD8+ T cell characteristics compared to CD4+ T cells. This lineage-level investigation involved both transcriptome and TCR features using HC samples. Linear mixed model analyses were performed conditioning the effects of donor batches and T cell-type batches (**Methods**).

In transcriptome analysis, 895 genes were differentially expressed (6.6% of the tested genes at FDR < 0.05, **Extended Figure 3a**). In alignment with their definitions, *CD8A*, *CD8B*, and *DBN1* genes exhibited distinctive expression in CD8+ T cells, whereas CD4+ T cells were distinguished by the gene expression of *CD4* and *FBLN7* (**Extended Figure 3b**).

In TCR feature analysis, we found significant associations in 108 TCR features, 68% of the tested features at FDR < 0.05: e.g., a higher frequency of acidic amino acids and a lower frequency of basic amino acids in CD8+ T cells than CD4+ T cells (**Extended Figure 3c**). We found that 34 out of 39 V gene and J gene differential usages in the β chains of CD8+ T cells were consistent with the findings of the previous report by Klarenbeek et al^1^ (Sign test P-value 2.4 × 10^-6^).

Additionally, the CDR3 regions of CD8+ T cells exhibited a higher frequency of acidic amino acids like aspartic acid (D) and glutamic acid (E), and a lower frequency of basic amino acids such as arginine (R) and lysine (K) (**Extended Figure 3d and 3e**). These results were consistent to the findings from previous studies ^1, 2^. These findings suggest a consistent and replicable distinction in TCR features between CD4+ and CD8+ T cells.

**HLA-TCR association analysis**

*HLA* alleles can be a strong confounding factor in TCR analysis since they correlate with both IMDs’ susceptibility and TCR repertoire sequences ^3, 4^. Therefore, we initially carried out association analyses between HLA alleles and the TCR features within each T cell-type, utilizing exclusively the samples from HC. As anticipated, we observed a higher number of Class 2 HLA alleles associated with TCR features of CD4+ T cell-types, and conversely, more associations with CD8+ T cells in the case of Class 1 HLA alleles (**Extended Figure 4**). On average, we have identified approximately 3.2 TCR sequence feature-associated HLA alleles and we have adjusted for their influence in TCR disease signature analysis (**Methods**).

**Intra-cell-type weighted variance partitioning analysis of transcriptome and TCR datasets**

To quantify the impact of IMDs in each T cell-type, we employed weighted variance partitioning analysis in each T cell-type using both the TCR feature and transcriptome datasets (**Methods**). The variation in disease conditions accounted for an average of 9.5% of the variance in the transcriptome and 2.1% of the variance in the TCR features (**Extended Figure** **5**). Among these cell-types, the transcriptome of Th1 exhibited the highest variability between disease conditions, accounting for 13.3% of the variance, showing the strong impact of IMDs on the heterogeneity of Th1 transcriptome.

Reference

1.	Klarenbeek, P.L. *et al.* Somatic Variation of T-Cell Receptor Genes Strongly Associate with HLA Class Restriction. *PLoS One* **10**, e0140815 (2015).

2.	Li, H.M. *et al.* TCRβ repertoire of CD4+ and CD8+ T cells is distinct in richness, distribution, and CDR3 amino acid composition. *J Leukoc Biol* **99**, 505-513 (2016).

3.	Ishigaki, K. *et al.* HLA autoimmune risk alleles restrict the hypervariable region of T cell receptors. *Nat Genet* **54**, 393-402 (2022).

4.	Nagafuchi, Y. *et al.* Control of naive and effector CD4 T cell receptor repertoires by rheumatoid-arthritis-risk HLA alleles. *J Autoimmun* **133**, 102907 (2022).
