## Supplementary figures for "T cell plasticity in systemic lupus erythematosus revealed by large-scale T cell receptor repertoire and transcriptome studies"

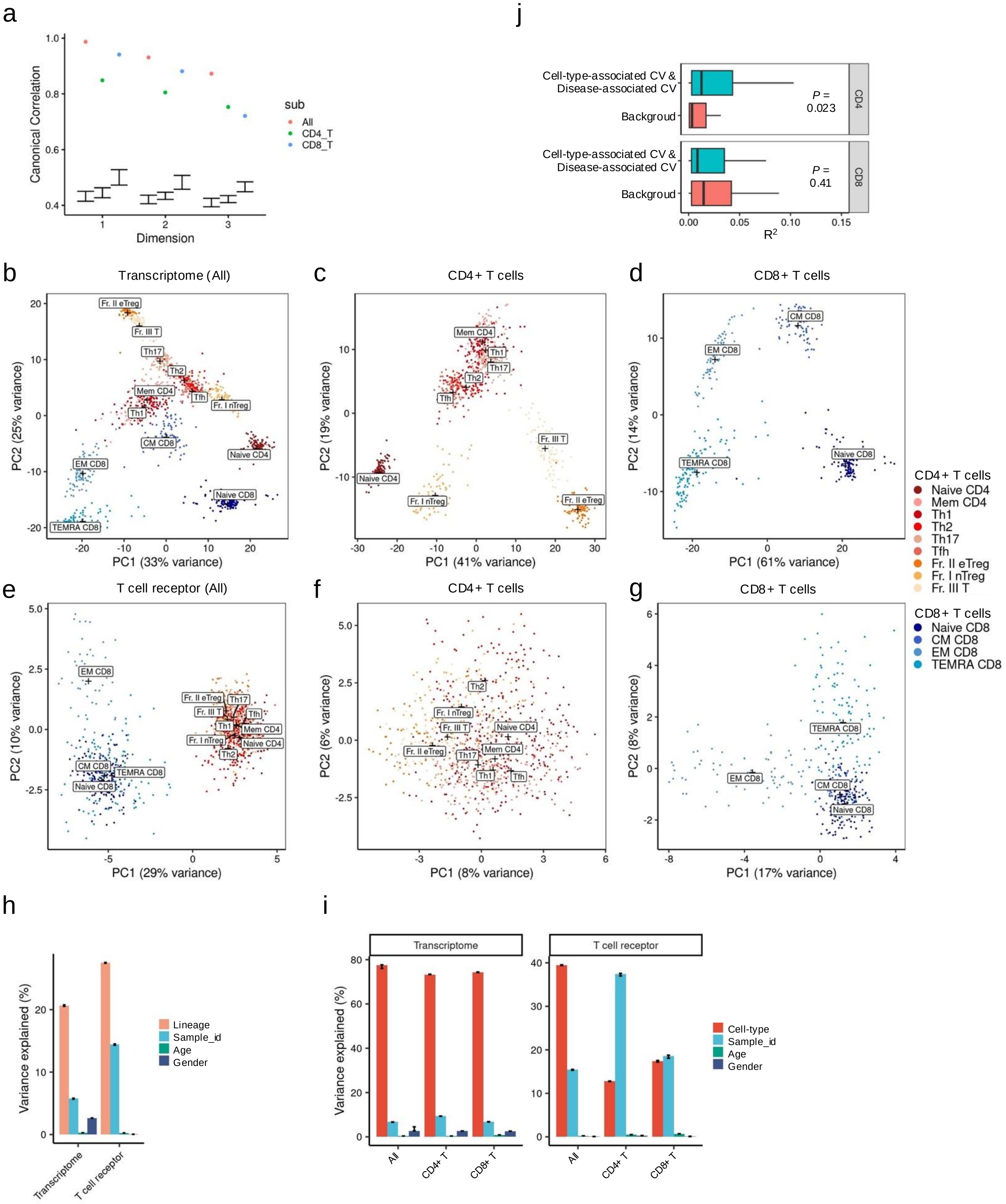

a
Canonical Correlation
Dimension
sub
All
CD4_T
CD8_T
j
Cell-type-associated CV &
Disease-associated CV
Backgroud
P =
0.023
0.41
CD4
CD8
R2
b
Transcriptome (All)
PC1 (33% variance)
PC2 (25% variance)
c
CD4+ T cells
PC1 (41% variance)
PC2 (19% variance)
d
CD8+ T cells
PC1 (61% variance)
PC2 (14% variance)
CD4+ T cells
Naive CD4
Mem CD4
Th1
Th2
Th17
Tfh
Fr. II eTreg
Fr. I nTreg
Fr. III T
e
T cell receptor (All)
PC1 (29% variance)
PC2 (10% variance)
f
CD4+ T cells
PC1 (8% variance)
PC2 (6% variance)
g
CD8+ T cells
PC1 (17% variance)
PC2 (8% variance)
CD8+ T cells
Naive CD8
CM CD8
EM CD8
TEMRA CD8
h
Variance explained (%)
Lineage
Sample_id
Age
Gender
Transcriptome
T cell receptor
i
Transcriptome
T cell receptor
Variance explained (%)
Cell-type
Sample_id
Age
Gender
All
CD4+ T
CD8+ T

**Supplementary Figure 1. Overview of the transcriptome and TCR data.**

(a) Canonical correlation of the first three TCR and transcriptome canonical variates (CVs). Error bars indicate the minimum and maximum canonical correlation observed across 100 permutation tests. (b-g) Principal component analysis (PCA) of transcriptome (b-d) and T cell receptor repertoire data (e-g). PCA was performed using all healthy control samples (b, e) and using CD4+ T cell (c,f) or CD8+ T cell (d,g) samples respectively. (h-i) Weighted variance partitioning analysis of the transcriptome and T cell receptor repertoire datasets. In g, the contribution of lineage (either CD4+ or CD8+) was analysed using all healthy control samples. In (h), the analysis was performed using all healthy control samples and using CD4+ T cell or CD8+ T cell samples respectively. Error bars indicate 95% confidence intervals from jackknife resampling. (j) Comparison of the association $R^2$ between loading vectors of cell-type-associated inter-cell-type CVs and disease-associated intra-cell-type CVs, and cell-type-non-associated inter-cell-type CVs and disease-non-associated intra-cell-type CVs (Background). The difference was tested with Mann-Whitney U test.

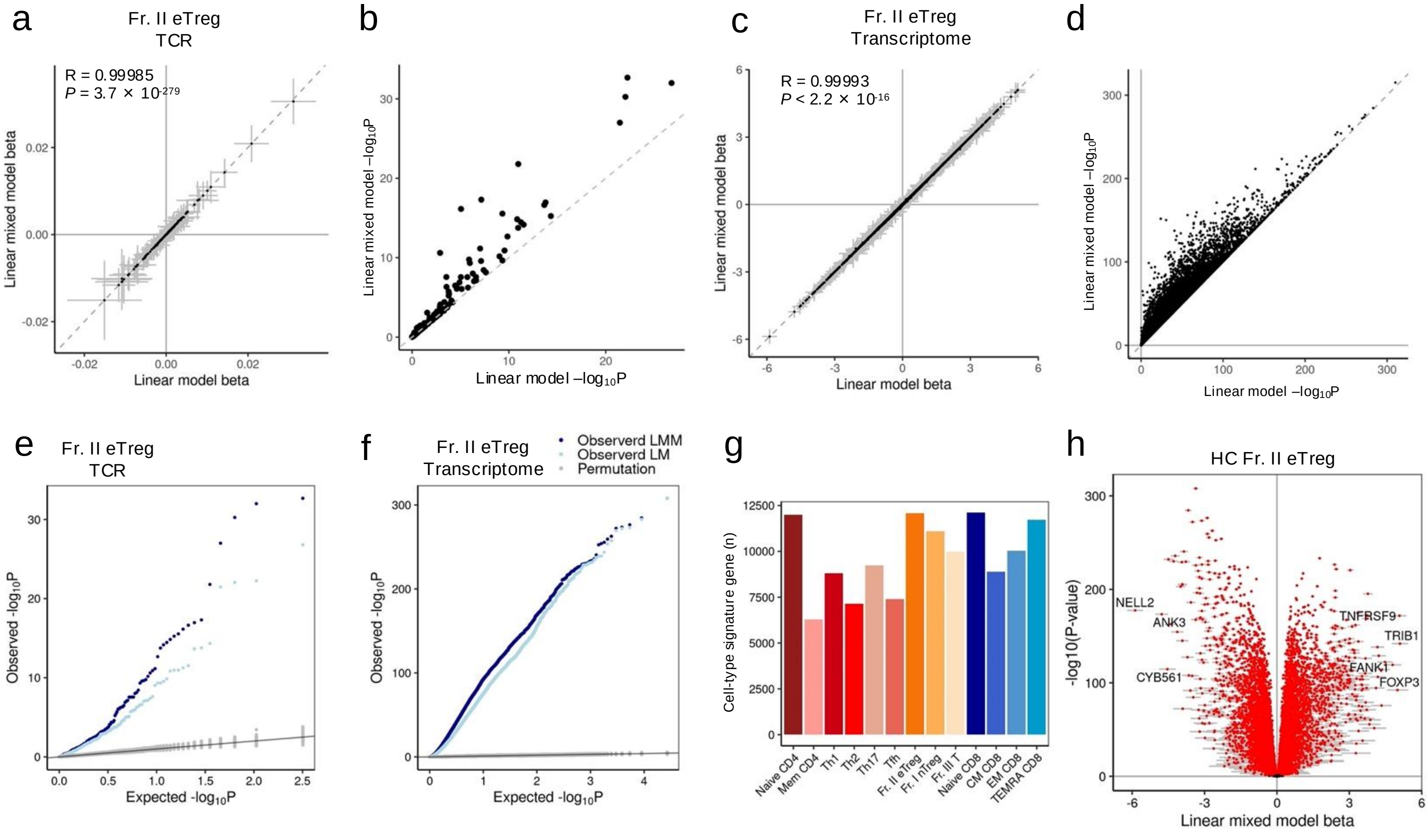


**Supplementary Figure 2. Comparison of the statistical models in cell-type signature analysis**

For the cell-type signature analysis of Fr. II eTreg, we examined the effect estimate beta and P-values by comparing the conventional linear model (LM) with the linear mixed model (LMM) in both the T cell receptor (TCR) analysis (a-b) and the transcriptome analysis (c-d). The analysis was restricted to samples from healthy controls. (e-f) Quantile-quantile plot comparing the observed P-value distribution of LMM, LM, and 1000 times permutation data in the Fr. II eTreg's TCR (e) and transcriptome (f) cell-type signature analysis. (g) The number of significant cell-type transcriptome signatures in T cell-types, determined at the LMM FDR < 0.05. (h) Transcriptome Fr. II eTreg signatures. Genes with FDR < 0.05 were noted with red color.

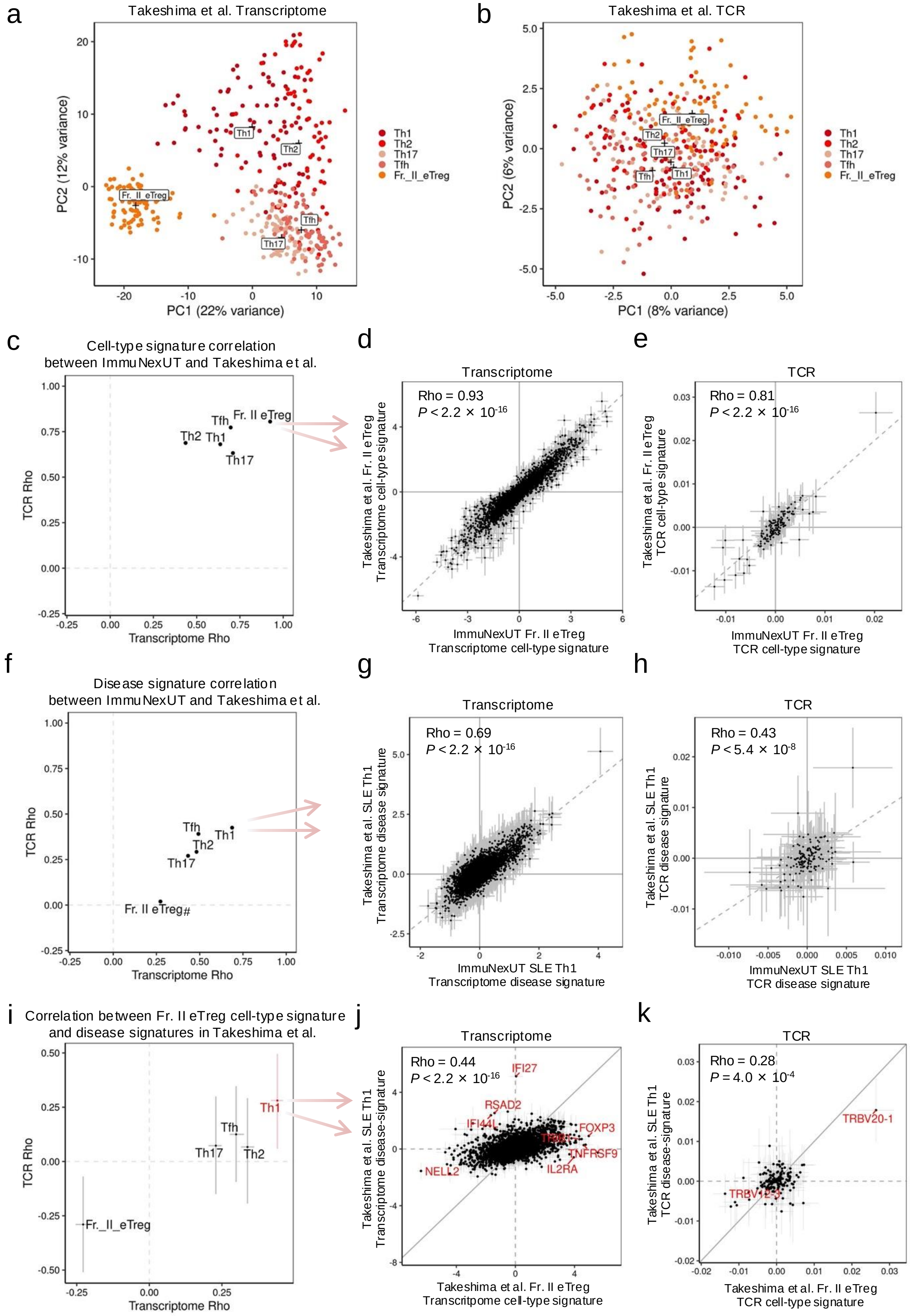
a
Takeshima et al. Transcriptome
PC1 (22% variance)
PC2 (12% variance)
Th1
Th2
Th17
Tfh
Fr._II_eTreg
b
Takeshima et al. TCR
PC1 (8% variance)
PC2 (6% variance)
Th1
Th2
Th17
Tfh
Fr._II_eTreg
c
Cell-type signature correlation
between ImmuNexUT and Takeshima et al.
TCR Rho
Transcriptome Rho
Tfh
Fr. II eTreg
Th2
Th1
Th17
d
Transcriptome
Rho = 0.93
P < 2.2 × 10-16
Takeshima et al. Fr. II eTreg
Transcriptome cell-type signature
ImmuNexUT Fr. II eTreg
Transcriptome cell-type signature
e
TCR
Rho = 0.81
P < 2.2 × 10-16
Takeshima et al. Fr. II eTreg
TCR cell-type signature
ImmuNexUT Fr. II eTreg
TCR cell-type signature
f
Disease signature correlation
between ImmuNexUT and Takeshima et al.
TCR Rho
Transcriptome Rho
Tfh
Th1
Th2
Th17
Fr. II eTreg#
g
Transcriptome
Rho = 0.69
P < 2.2 × 10-16
Takeshima et al. SLE Th1
Transcriptome disease signature
ImmuNexUT SLE Th1
Transcriptome disease signature
h
TCR
Rho = 0.43
P < 5.4 × 10-8
Takeshima et al. SLE Th1
TCR disease signature
ImmuNexUT SLE Th1
TCR disease signature
i
Correlation between Fr. II eTreg cell-type signature
and disease signatures in Takeshima et al.
TCR Rho
Transcriptome Rho
Th1
Tfh
Th17
Th2
Fr._II_eTreg
j
Transcriptome
Rho = 0.44
P < 2.2 × 10-16
IFI27
RSAD2
IFI44L
FOXP3
TNFRSF9
IL2RA
NELL2
Takeshima et al. SLE Th1
Transcriptome disease-signature
Takeshima et al. Fr. II eTreg
Transcritpome cell-type signature
k
TCR
Rho = 0.28
P = 4.0 × 10-4
TRBV20-1
Takeshima et al. SLE Th1
TCR disease-signature
Takeshima et al. Fr. II eTreg
TCR cell-type signature

**Supplementary Figure 3. Validation analysis of Takeshima et al. RNA-seq data**

(a, b) Principal component analysis (PCA) of transcriptome (a) and T cell receptor (TCR) features (b) of Takeshima et al. PCA was performed using healthy control samples. (c, f) Comparison of Spearman's correlation coefficients in CD4 T cell-types between Transcriptome and TCR cell-type signature analysis (c) and disease signature analysis (f). X axis displayed the Spearman's correlation coefficients between the transcriptome Fr. II eTreg cell-type signature betas of ImmuNexUT discovery dataset and Takeshima et al. validation dataset analysis. Y axis represented the TCR counterparts. In (f), # indicated P-values > 0.05/5 in Spearman's correlation analysis of TCR signatures. (d, e) Representative correlation plots of Fr. II eTreg cell-type signatues by transcriptome (d) and TCR analysis (e). (g, h) Representative correlation plots of SLE Th1 disease signatues by transcriptome (g) and TCR analysis (h). (i) Comparison of Spearman's correlation coefficients between cell-type and disease signatures in Takeshima et al dataset. X axis displayed the Spearman's correlation coefficients with the 99% confidence interval, focusing on the correlation between the transcriptome Fr. II eTreg cell-type signature and SLE disease signature in CD4 T cell-types. Y axis represented the TCR counterparts. (j-k) The correlation plots between SLE Th1 disease signature and Fr. II eTreg cell-type signature by Takeshima et al validation cohort transcriptional (j) and TCR (k) data.

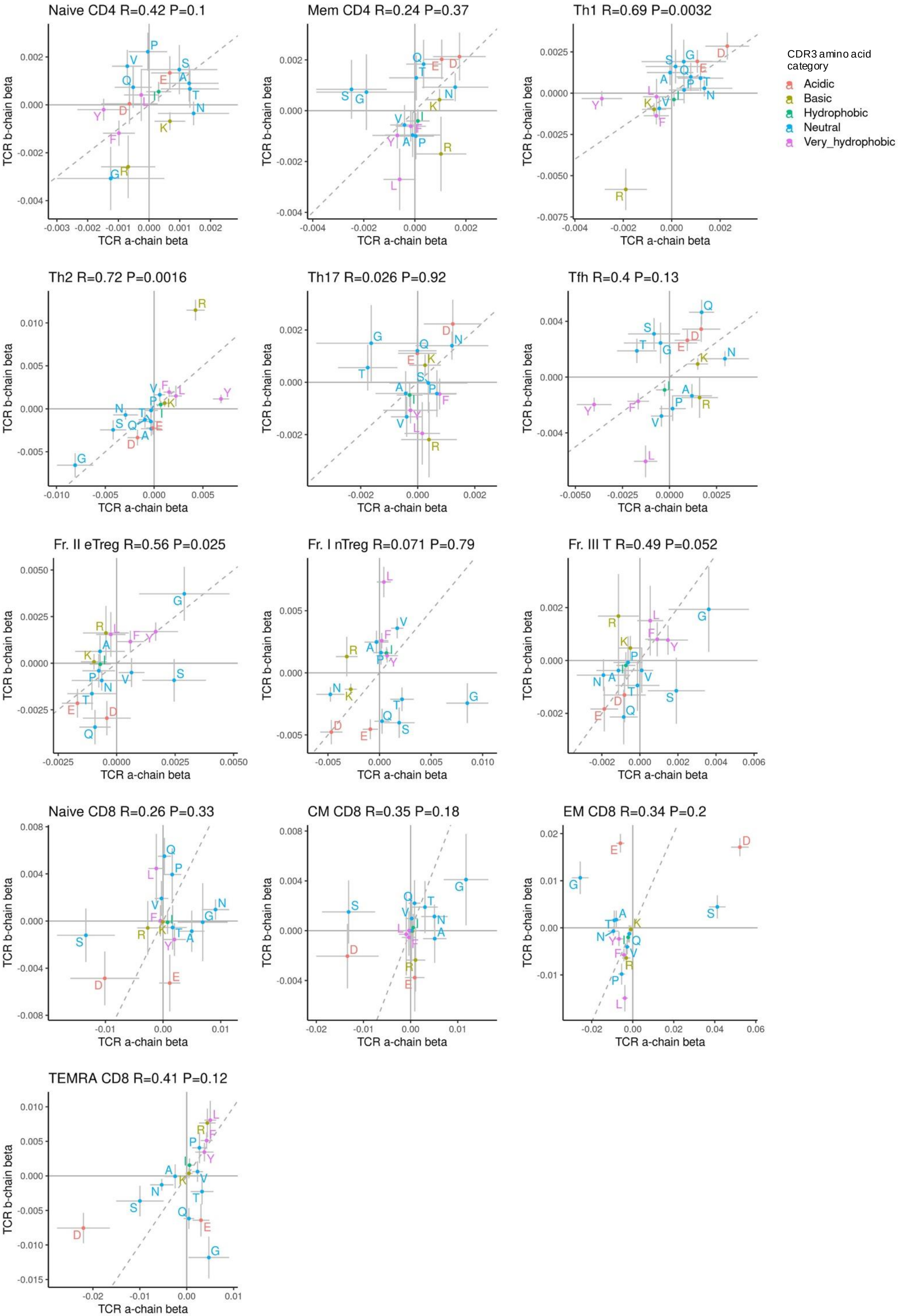
Naive CD4 R=0.42 P=0.1
Mem CD4 R=0.24 P=0.37
Th1 R=0.69 P=0.0032
Th2 R=0.72 P=0.0016
Th17 R=0.026 P=0.92
Tfh R=0.4 P=0.13
Fr. II eTreg R=0.56 P=0.025
Fr. I nTreg R=0.071 P=0.79
Fr. III T R=0.49 P=0.052
Naive CD8 R=0.26 P=0.33
CM CD8 R=0.35 P=0.18
EM CD8 R=0.34 P=0.2
TEMRA CD8 R=0.41 P=0.12
TCR a-chain beta
TCR b-chain beta
CDR3 amino acid category
Acidic
Basic
Hydrophobic
Neutral
Very_hydrophobic

**Supplementary Figure 4. T cell receptor CDR3 amino acid usage signatures of T cell-types.**

In the analysis of CDR3 amino acid usages of T cell receptor cell-type signatures, amino acids were categorized based on their physicochemical features, and the signature betas were compared between TCR α and β chains with Pearson Correlation Coefficient.

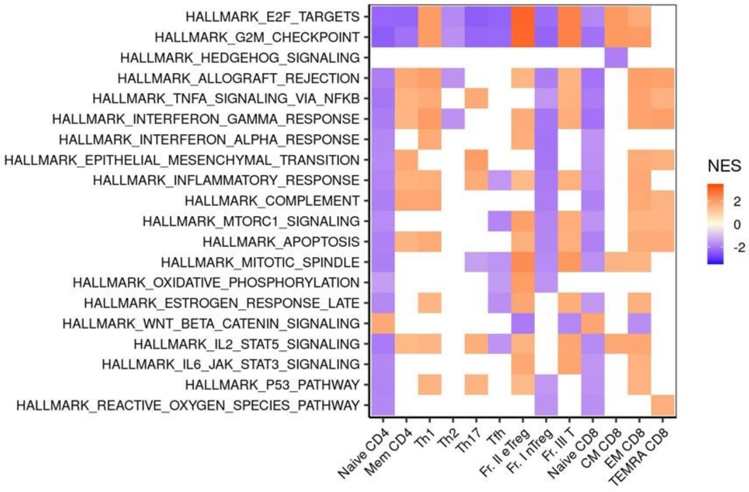


**Supplementary Figure 5. Pathway analysis of transcriptome cell-type signature.**

Gene Set Enrich Analysis of transcriptome cell-type signatures. Pathways with |NES| > 1.8 in at least in one of the cell-types were visualized.
NES, Normalized enrichment score.

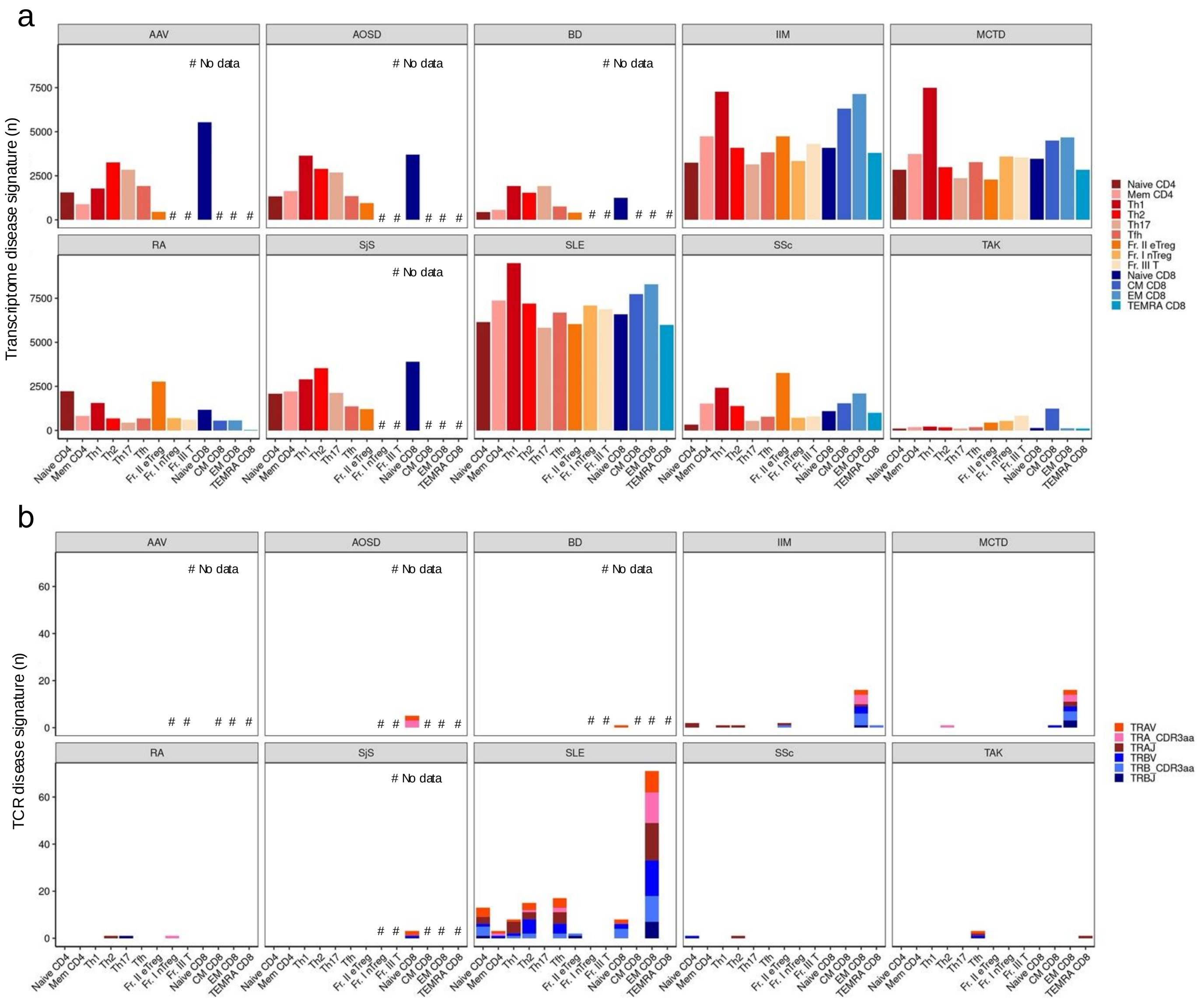


**Supplementary Figure 6. Disease signature analysis.**

(a-b) The number of transcriptome (a) and T cell receptor (TCR) features (b) disease signature at FDR < 0.05. # indicated the five T cell-types that were not included in the study.
SLE, Systemic lupus erythematosus; SSc, Systemic sclerosis; IIM, Idiopathic inflammatory myopathy; RA, Rheumatoid arthritis; AAV, ANCA-associated vasculitis; BD, Behçet's disease; MCTD, Mixed connective tissue disease; AOSD, Adult-onset Still’s disease; SS, Sjögren’s syndrome; TAK, Takayasu arteritis.

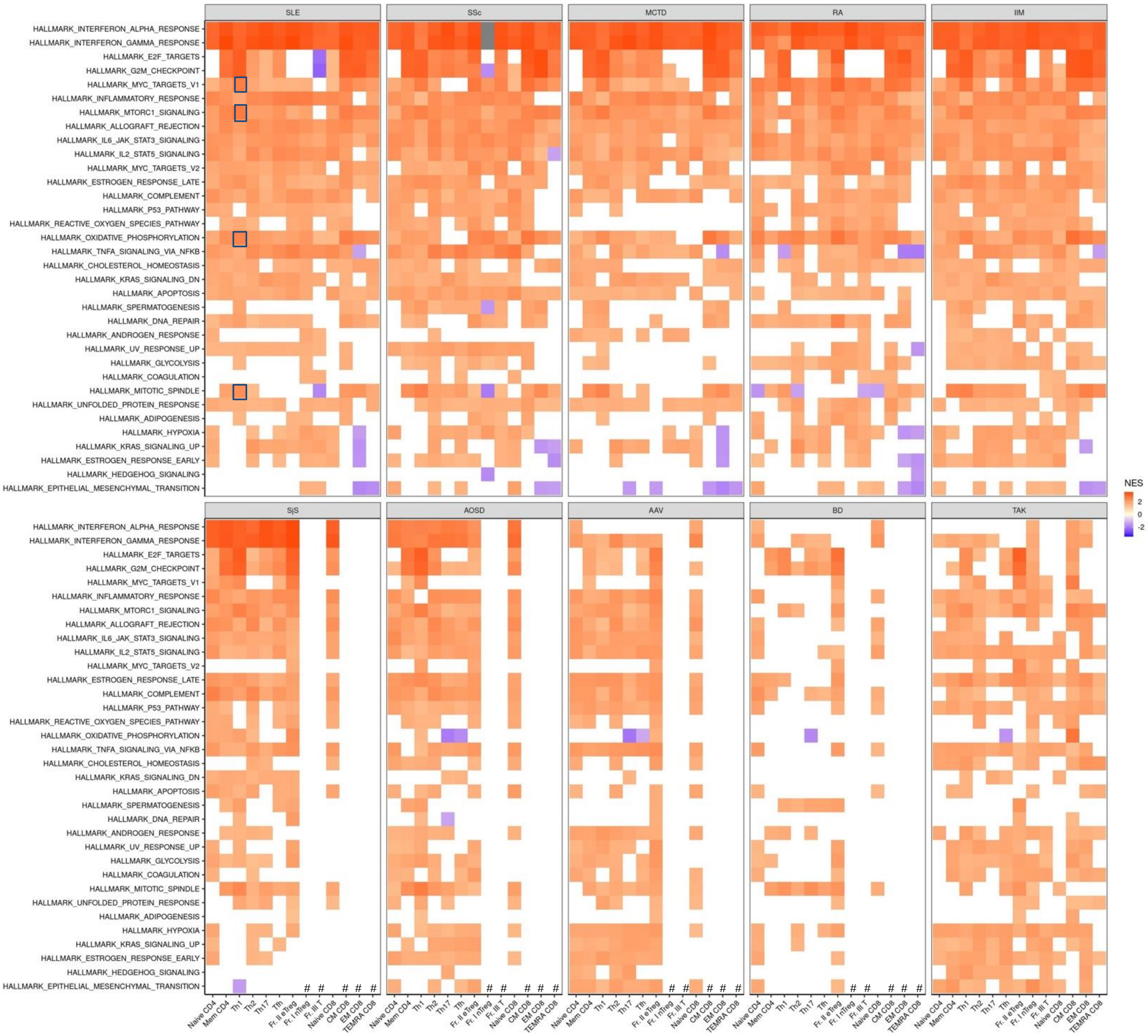


**Supplementary Figure 7. Pathway analysis of disease gene signature.**

Gene Set Enrich Analysis of transcriptome disease signature in immune mediated diseases. Pathways with |NES| > 2 in at least one of the cell-types were visualized. # indicated the five T cell-types that were not included in the study in ImmuNexUT phase 1.

NES, Normalized enrichment score; SLE, Systemic lupus erythematosus; SSc, Systemic sclerosis; IIM, Idiopathic inflammatory myopathy; RA, Rheumatoid arthritis; AAV, ANCA-associated vasculitis; BD, Behçet's disease; MCTD, Mixed connective tissue disease; AOSD, Adult-onset Still’s disease; SS, Sjögren’s syndrome; TAK, Takayasu arteritis.

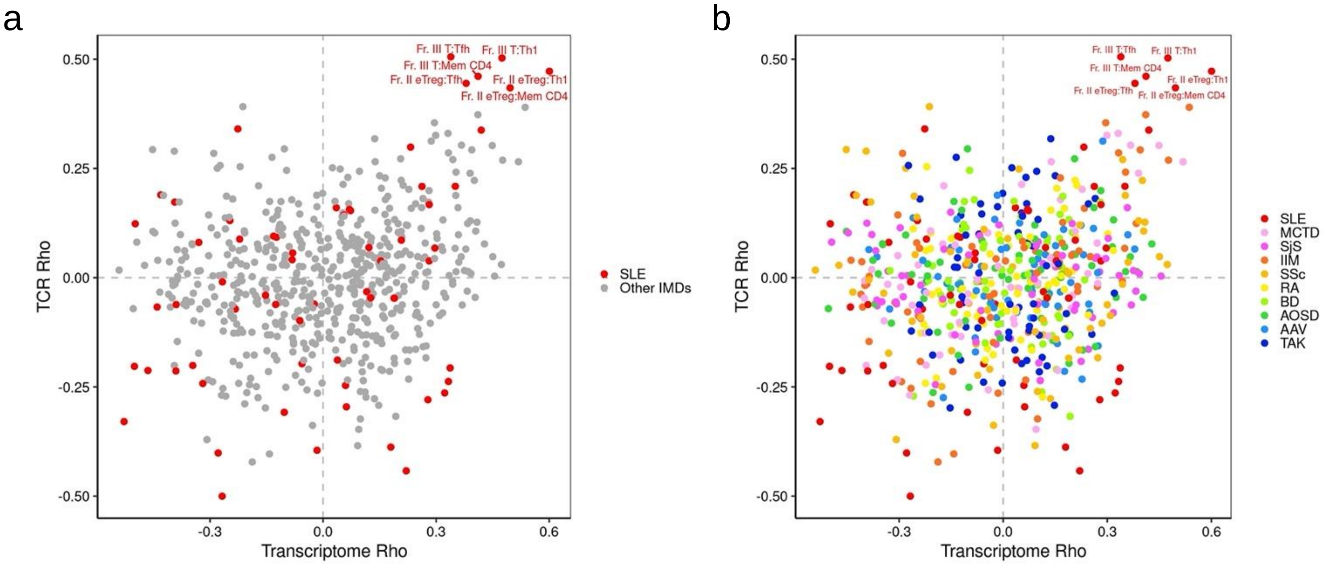


**Supplementary Figure 8. Correlation analysis in all combination of cell-type signatures and disease signatures.**

(a-b) Comparison of Spearman's correlation coefficients between Transcriptome and T cell receptor (TCR) analysis. X axis displayed the Spearman's correlation coefficients between the transcriptome cell-type signature and disease signature in CD4+ T cell-types. Y axis represented the TCR counterparts. In (a), red color indicated SLE and in (b) color represented each immune-mediated diseases. The label "Fr. II eTreg:Th1" represented the correlation coefficients between cell-type signature of Fr. II eTreg and disease signature of Th1.

SLE, Systemic lupus erythematosus; SSc, Systemic sclerosis; IIM, Idiopathic inflammatory myopathy; RA, Rheumatoid arthritis; AAV, ANCA-associated vasculitis; BD, Behçet's disease; MCTD, Mixed connective tissue disease; AOSD, Adult-onset Still's disease; SS, Sjögren's syndrome; TAK, Takayasu arteritis.

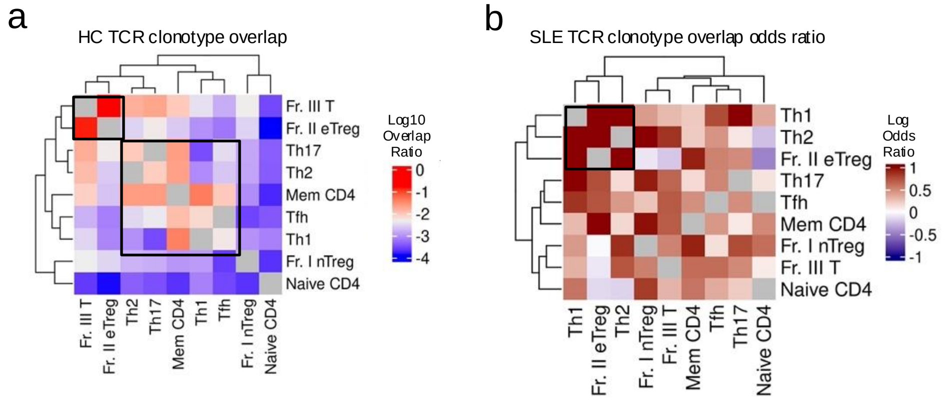


**Supplementary Figure 9. TCR clonotype overlap analysis.**

(a) A heatmap showing the TCR clonotype overlap ratio across CD4+ T cell-types. The order of T cell-type in row and column were based on the hierarchical clustering. We analyzed only the TCR clonotype overlap between healthy control (HC) samples from ImmuNexUT Phase 2. (b) A heatmap showing the odds ratio of TCR clonotype overlap in SLE in contrast to healthy controls in all combination of CD4+ T cell-types based on logistic mixed effect analysis. The order of T cell-type in row and column were based on the hierarchical clustering.

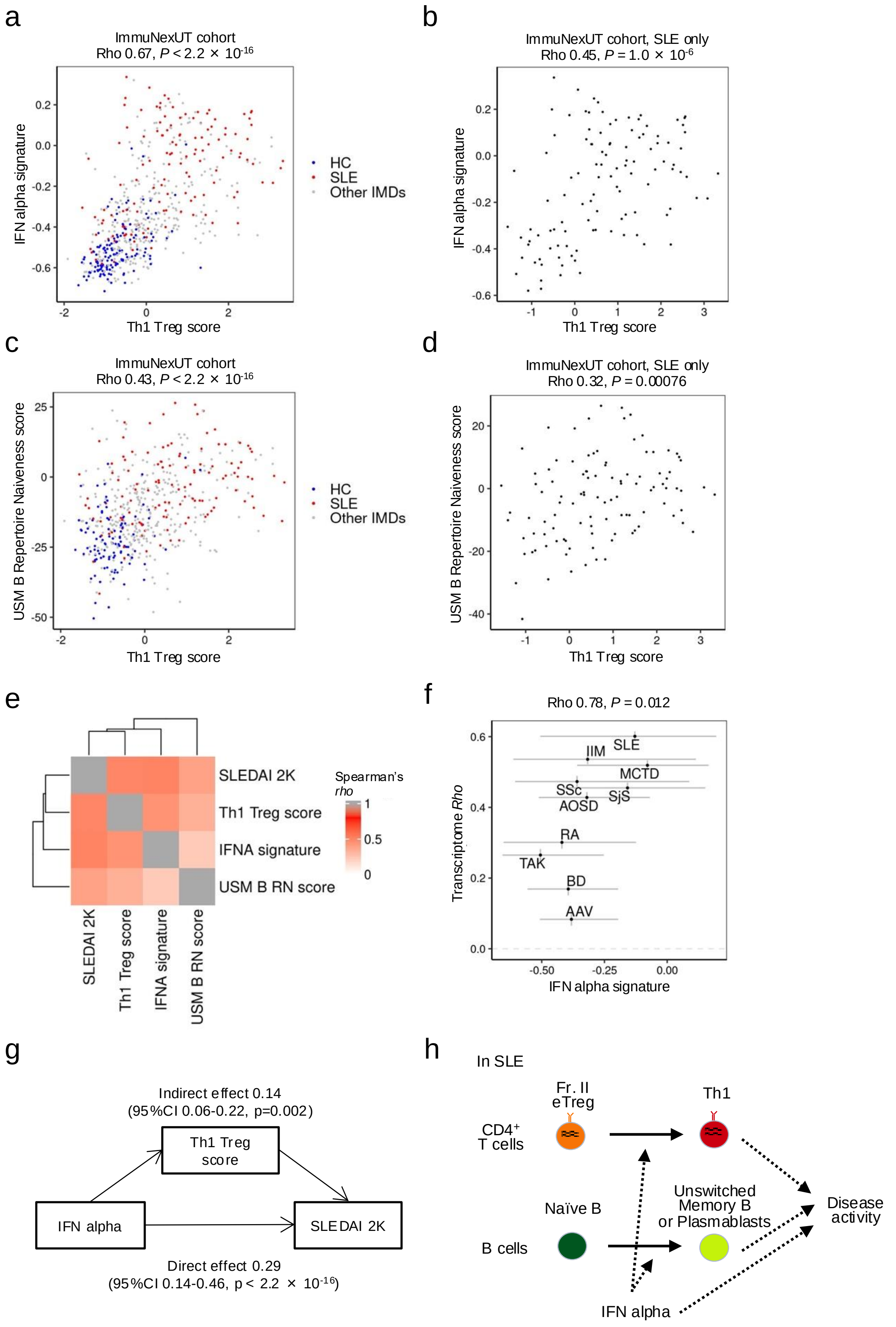
a
ImmuNexUT cohort
Rho 0.67, P < 2.2 × 10-16
IFN alpha signature
Th1 Treg score
HC
SLE
Other IMDs
b
ImmuNexUT cohort, SLE only
Rho 0.45, P = 1.0 × 10-6
IFN alpha signature
Th1 Treg score
c
ImmuNexUT cohort
Rho 0.43, P < 2.2 × 10-16
USM B Repertoire Naiveness score
Th1 Treg score
HC
SLE
Other IMDs
d
ImmuNexUT cohort, SLE only
Rho 0.32, P = 0.00076
USM B Repertoire Naiveness score
Th1 Treg score
e
SLEDAI 2K
Th1 Treg score
IFNA signature
USM B RN score
Spearman's rho
f
Rho 0.78, P = 0.012
Transcriptome Rho
IFN alpha signature
SLE
IIM
MCTD
SSc
SjS
AOSD
RA
TAK
BD
AAV
g
Indirect effect 0.14
(95%CI 0.06-0.22, p=0.002)
Th1 Treg score
IFN alpha
SLEDAI 2K
Direct effect 0.29
(95%CI 0.14-0.46, p < 2.2 × 10-16)
h
In SLE
CD4+ T cells
Fr. II eTreg
Th1
B cells
Naïve B
Unswitched Memory B or Plasmablasts
Disease activity
IFN alpha

**Supplementary Figure 10. Correlation of Th1 Treg score, Interferion signature, and B cell repertoire naiveness score.**

(a-d) Scatter plots illustrating the correlation between the Treg score and Interferon (IFN) alpha signature in Th1 (a-b) or between the Treg score in Th1 and the Repertoire Naiveness score in unswitched memory (USM) B cells from Ota et al. (c-d). In (a) and (c), the Spearman's correlation is depicted across the entire ImmuNexUT cohort, and in (b) and (d), stratified analysis of SLE patients is presented. (e) Correlation heatmap between SLEDAI-2K, Th1 Treg score, IFN alpha signature, and USM B RN score in SLE patients. (f) Scatter plots illustrating the correlation between the mean IFN-α signature in Th1 cells and the Spearman's correlation coefficients between the transcriptomic Fr. II eTreg cell-type signature and the disease signature of IMDs in Th1 cells. The error bars on the x-axis represent the 5th and 95th percentile distribution of the IFN-α signature, while the error bars on the y-axis represent the 95% confidence intervals. (g) Mediation model representing the relationships between IFN alpha signature, Th1 Treg score, and SLEDAI 2K. (h) Schematic representation of the CD4+ T cell and B cell plasticity and IFN alpha in SLE.
