## Extended figures for "T cell plasticity in systemic lupus erythematosus revealed by large-scale T cell receptor repertoire and transcriptome studies"

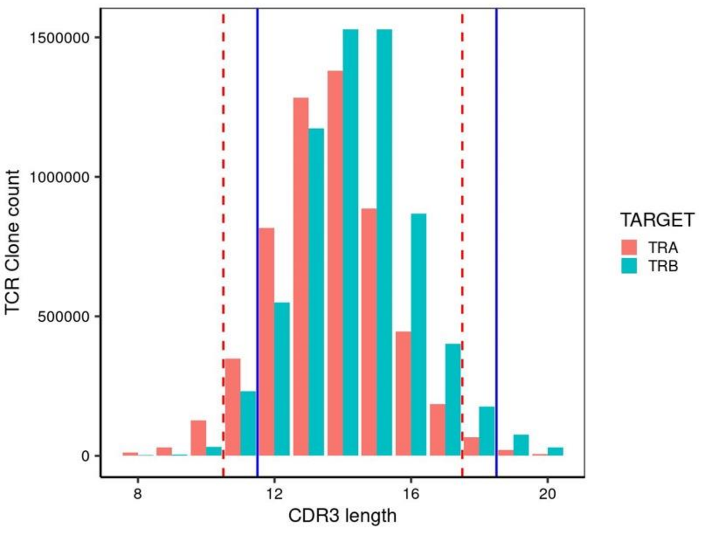


**Extended Figure 1. CDR3 amino acids length in TCR α-chains and β chains.**

The distribution of CDR3 amino acids length in TCR α-chains and β chains of the ImmuNexUT discovery cohort. In the CDR3 amino acid usage analysis, we included only the TCR clones falling within the specified window of CDR3 length (11-17 for TCR α-chain indicated by the dashed red line and 12-18 for TCR β-chain denoted by blue lines). We computed the average frequency of CDR3 amino acid usages by considering the middle amino acid positions (IMGT P108-P112) in the CDR3 region of these samples.

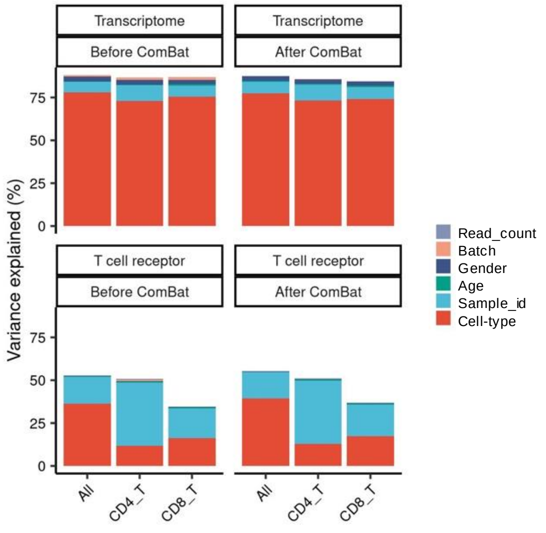


**Extended Figure 2. Weighted variance partitioning analyses of the transcriptome and T cell receptor repertoire datasets.**

Weighted variance partitioning analysis of the transcriptome and T cell receptor repertoire datasets before and after ComBat procedure. Linear mixed model formula was applied for both the data prior to and after implementing the ComBat procedure to adjust for the known study phase batch effects (Methods). The Batch represents two study phase batches. The analysis was performed using all healthy control samples and using CD4+ T cell or CD8+ T cell samples respectively.

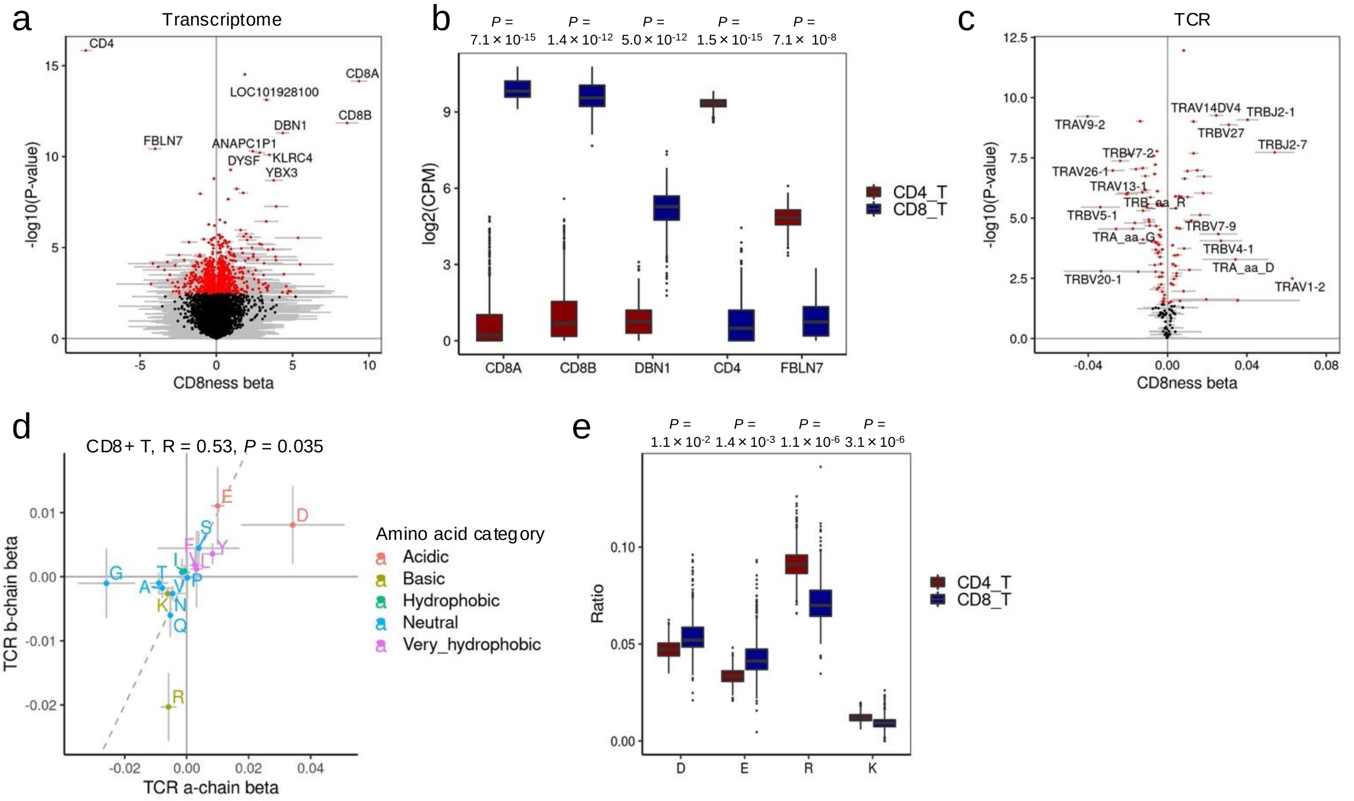


**Extended Figure 3. CD8+ T cell signature analysis**

(a,c) Transcriptome and T cell receptor (TCR) CD8+ T cell signatures. Genes with FDR<0.05 were noted with red color. Grey bar in each dot indicated ±2 SE ranges. (b) Representative boxplots of CD8+ and CD4+ T cell signature genes. P-values were derived from CD8 T cell signature analysis. (d) TCR CDR3 amino acid usage signatures in CD8+ T cells. Amino acids were categorized based on their physicochemical features, and the signature betas were compared between TCR α and β chains with Pearson Correlation Coefficient. (e) Representative boxplots of CD8+ and CD4+ TCR CDR3 amino acid usage in β chains. P-values were derived from cell-type signature analysis.

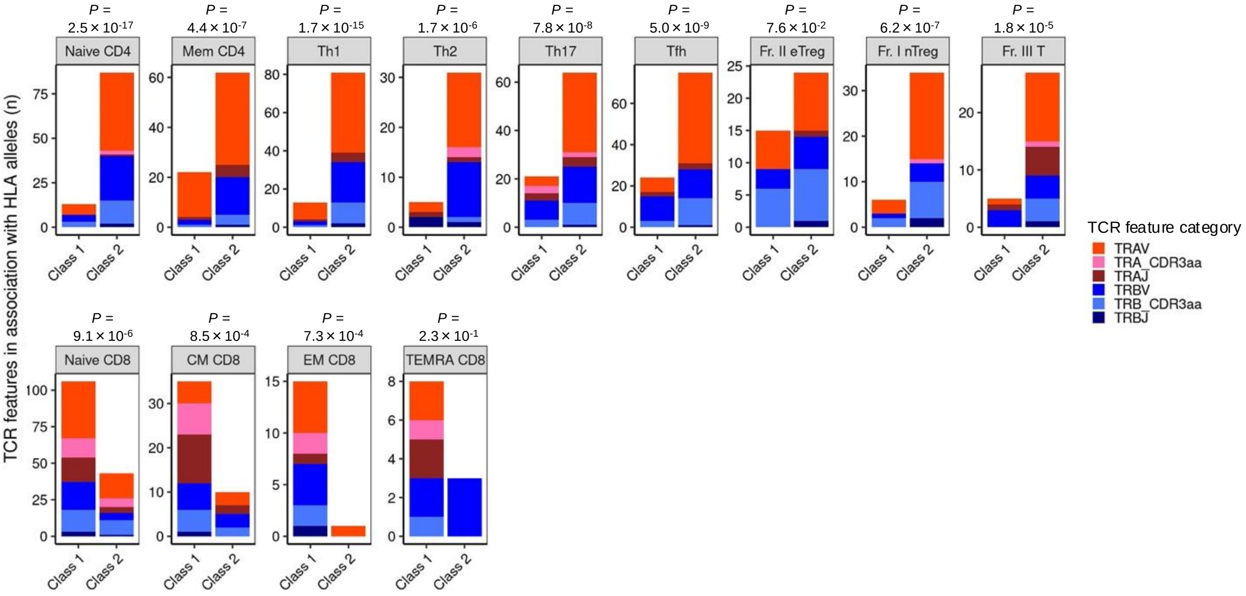


**Extended Figure 4. Association of HLA and TCR features**

The number of T cell receptor (TCR) features linked to Class 1 and Class 2 HLA alleles. P-values were computed using Fisher's exact test. TCR features were classified based on distinctions between TCR α and β chains and variations in the category (V and J gene usages and CDR3 amino acid usages).

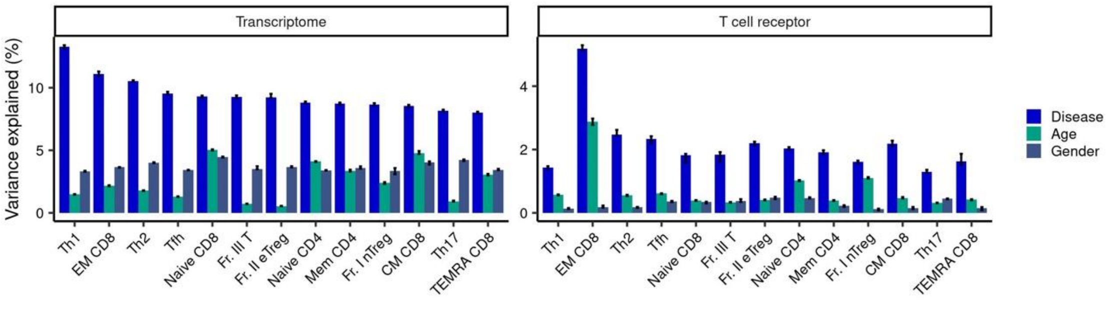


**Extended Figure 5. Transcriptome and TCR variation in immune mediated diseases.**

Intra-cell-type weighted variance partitioning analysis of the transcriptome and T cell receptor (TCR) datasets. The analysis was performed in each T cell-type respectively. The order of T cell-type is the same and based on the order of the explained variance by the disease in transcriptome analysis. Error bars indicate 95% confidence intervals from jackknife resampling.
